# Preventing tuberculosis with community-based care in an HIV-endemic setting: a modeling analysis

**DOI:** 10.1101/2023.08.21.23294380

**Authors:** Jennifer M. Ross, Chelsea Greene, Cara J. Bayer, David W. Dowdy, Alastair van Heerden, Jesse Heitner, Darcy W. Rao, D. Allen Roberts, Adrienne E. Shapiro, Zelda B. Zabinsky, Ruanne V. Barnabas

## Abstract

**Introduction:** Antiretroviral therapy (ART) and TB preventive treatment (TPT) both prevent tuberculosis (TB) disease and deaths among people living with HIV. Differentiated care models, including community-based care, can increase uptake of ART and TPT to prevent TB in settings with a high burden of HIV-associated TB, particularly among men.

**Methods:** We developed a gender-stratified dynamic model of TB and HIV transmission and disease progression among 100,000 adults ages 15-59 in KwaZulu-Natal, South Africa. We drew model parameters from a community-based ART initiation and resupply trial in sub-Saharan Africa (Delivery Optimization for Antiretroviral Therapy, DO ART) and other scientific literature. We simulated the impacts of community-based ART and TPT care programs during 2018-2027, assuming that community-based ART and TPT care were scaled up to similar levels as in the DO ART trial (i.e., ART coverage increasing from 49% to 82% among men and from 69% to 83% among women) and sustained for ten years. We projected the number of TB cases, deaths, and disability-adjusted life years (DALYs) averted relative to standard, clinic-based care. We calculated program costs and incremental cost-effectiveness ratios from the provider perspective.

**Results:** If community-based ART care could be implemented with similar effectiveness to the DO ART trial, increased ART coverage could reduce TB incidence by 27.0% (range 21.3% - 34.1%) and TB mortality by 36.0% (range 26.9% - 43.8%) after ten years. Increasing both ART and TPT uptake through community-based ART with TPT care could reduce TB incidence by 29.7% (range 23.9% - 36.0%) and TB mortality by 36.0% (range 26.9% - 43.8%). Community-based ART with TPT care reduced gender disparities in TB mortality rates by reducing TB mortality among men by a projected 39.8% (range 32.2% - 46.3%) and by 30.9% (range 25.3% - 36.5%) among women. Over ten years, the mean cost per DALY averted by community-based ART with TPT care was $846 USD (range $709 - $1,012).

**Conclusions:** By substantially increasing coverage of ART and TPT, community-based care for people living with HIV could reduce TB incidence and mortality in settings with high burdens of HIV-associated TB and reduce TB gender disparities.

## Introduction

Tuberculosis (TB) is the leading cause of death globally among people living with HIV (PLWH) [1]. HIV infection markedly increases the risk of progression to active TB disease [2]. The burden of HIV-associated TB is particularly high in South Africa, where more than 50% of people with incident TB in 2021 also had HIV, compared to a global mean of 6.7% [1]. Despite frequent co-occurrence, TB and HIV exhibit different gender disparity patterns in South Africa, where TB prevalence nationally is higher among men compared to higher HIV prevalence among women [3,4]. Multiple factors likely contribute to greater TB burden among men, including gender differences in accessing health care, gender-specific contact patterns, and higher prevalence of exposures that increase risk for TB infection and/or progression (e.g., mining, incarceration, use of alcohol, illicit substances, and tobacco) [5–7]. Additionally, gender disparities in HIV outcomes include lower levels of HIV viral suppression among men than women living with HIV across nearly all regions globally and in South Africa [8]. The higher detectable viral loads in men contributes in part to higher incidence among women [1].

TB preventive treatment (TPT) when given with antiretroviral therapy (ART), reduces the risk of TB by approximately one third [9–11]. Between 2018 and 2020, over six million PLWH accessed TPT globally, exceeding the target set at the 2018 UN High-Level Meeting on Tuberculosis. However, disruptions in care during the COVID-19 pandemic were associated with a 21% decline in the number of people taking TPT between 2019 and 2020 [1]. While global TPT figures are not disaggregated by gender, some studies conducted between 2020 and 2021 in sub-Saharan Africa found lower rates of TPT initiation and completion among men than among women [12,13], while others did not find differences [14,15].

Differentiated models of care tailor health care delivery to client needs and often extend care beyond traditional facility settings and involve an additional cadre of health workers [16]. Differentiated community-based care models have been effective and cost-effective interventions for HIV and TB care in sub-Saharan Africa [17], and can improve healthcare by caring for PLWH outside of health facilities. Recently, the Delivery Optimization for Antiretroviral Therapy (DO ART) household-randomized trial of community-based ART initiation and treatment in South Africa and Uganda demonstrated that community-based care increases the proportion of PLWH who achieve viral suppression, particularly among men [18]. This overcame a gender disparity in viral suppression that was observed in facility-based care models. At South African DO ART sites, asymptomatic participants in the community-based care group without contraindications to TPT were offered TPT starting one month after ART initiation, while the standard clinic group received TPT per routine clinic procedures. TPT uptake was higher among participants who received community-based care than participants who received facility-based care [19]. A recent modelling analysis concluded that community-based HIV treatment was cost-effective in preventing death and disability due to HIV [20]. However, the longer-term impact of increased ART and TPT uptake on incident TB and TB deaths were not quantified among trial participants, nor were TB incidence or deaths quantified among community members who were not in the trial.

In this modeling study, we aim to quantify the health impact and cost-effectiveness of extending community-based ART and TPT care to population scale in a TB-HIV high-burden setting of KwaZulu-Natal, South Africa, including analysis of gender disparities in health outcomes. In KwaZulu-Natal, where approximately 70% of people who develop TB also have HIV [21], we hypothesize that reaching the ART and TPT coverage levels achieved in the DO ART trial among all PLWH would substantially reduce TB disease and deaths and reduce gender disparities in TB.

## Methods

### Study Design and Setting

We developed a dynamic transmission model to simulate TB and HIV outcomes in KwaZulu-Natal, South Africa. We modeled three care delivery programs, including standard facility-based ART and TPT care (Program 1), community-based ART while assuming the number of individuals initiating TPT occurs at the facility-based care rate (Program 2), and community-based ART with TPT care (Program 3). Program 2 was not implemented in DO ART, but is modeled to evaluate the independent effects of ART and TPT. We executed the model over a 10-year intervention period to project health and economic outcomes – including TB incidence, TB mortality, disability-adjusted life years (DALYs), and costs – for each care delivery program.

### Dynamic Transmission Model

The dynamic transmission model is stratified by TB stage, HIV stage, TB and HIV treatment status, TB drug resistance status, and gender. We simulate a population of 100,000 persons that represent the adult population (ages 15-59) of KwaZulu-Natal, South Africa. The population moves through the compartments at transition rates reflecting TB and HIV infection, disease progression, treatment and recovery from TB, death, and aging into and out of the system (Fig. 1). The system of ordinary differential equations is described in the appendix, and is solved in R using the deSolve package in time steps of one month [22]. Model code is available in a public repository (https://github.com/cgreene3/epi_model_HIV_TB_KZN_SA). The model includes TB compartments for uninfected, latent TB infection (LTBI), active TB disease, and recovered/treated populations, with LTBI divided into recently infected (within two years), and remotely infected (more than two years). Those with recent infections have a higher probability of progression to active TB [23]. The model considers drug-susceptible TB (DS-TB) and multi-drug-resistant TB (MDR-TB) infections. We define MDR-TB as resistant to isoniazid and rifampicin and assume that individuals taking TPT are not protected against acquiring an MDR-TB infection.

**Figure 1.**
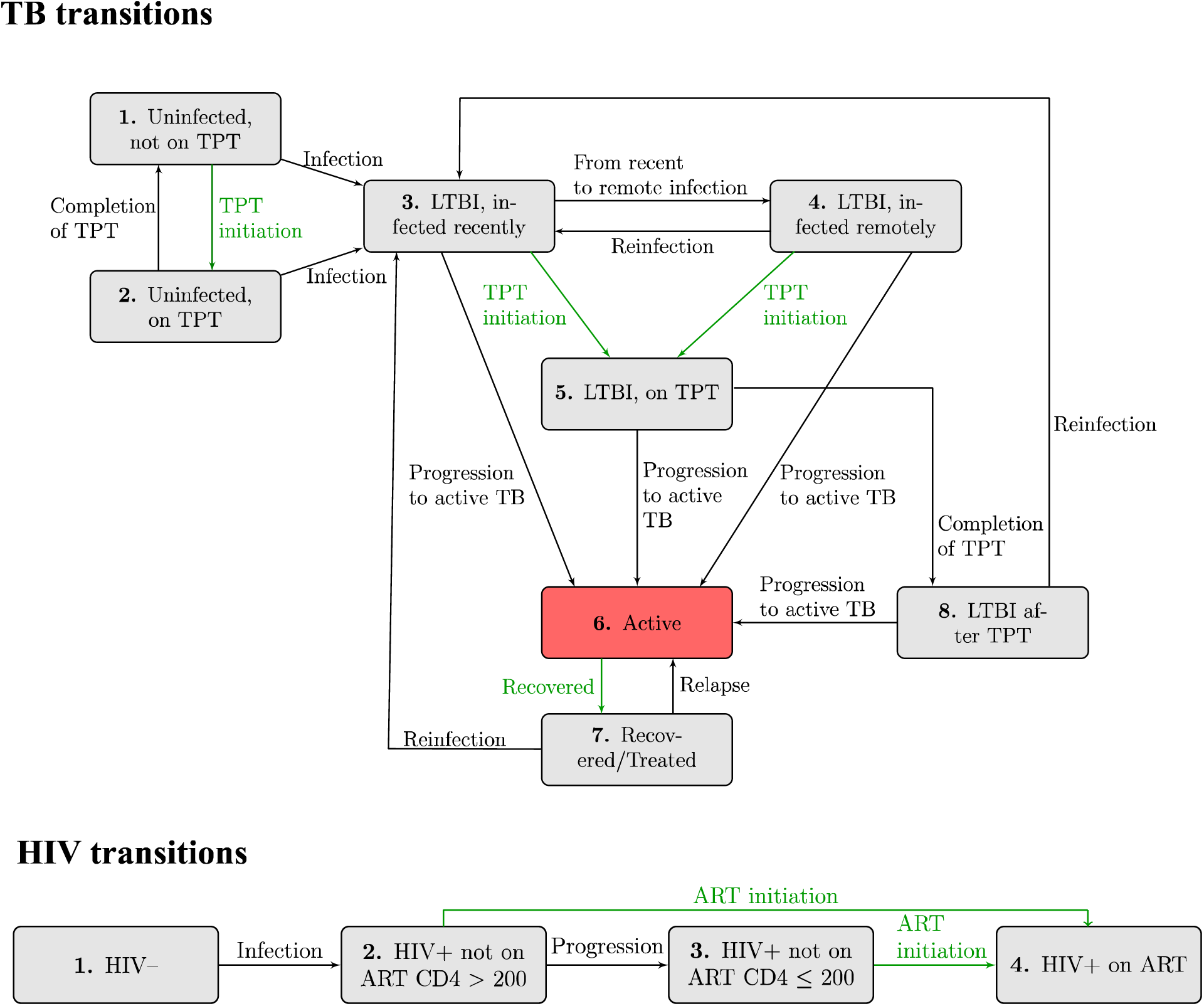
Illustration of TB and HIV dynamic transmission model. Rates of flow between each compartment are governed by differential equations, as described in the appendix. Although not visualized here, each TB and HIV compartment is stratified across two TB drug-resistance categories and two genders. The latent TB infection (LTBI) compartment is distinguished by those infected within two years, considered recent, and more than two years, considered remote. The rate of TB preventive treatment (TPT) and antiretroviral therapy (ART) initiations (highlighted in green) are directly impacted by care delivery programs (Table 2). ART coverage is used to calculate ART initiation rates so that the proportion of PLWH on ART by gender corresponds to ART coverage assumptions. The active TB compartment is highlighted in red to emphasize the compartment capturing incident TB.

The model includes HIV compartments for people living without HIV, PLWH with CD4>200, with CD4≤200, and taking ART (any CD4 count). Annual gender-specific HIV incidence estimates for KwaZulu-Natal, South Africa are incorporated from the Global Burden of Disease, Risk Factors, and Injuries Study 2019 (GBD 2019) [24] and the Data-driven Recommendations for Interventions against Viral Infection model [25], as described in the appendix. Model parameters that differ by HIV characteristics include the relative transmissibility of TB [26, 27], risk of progression to active TB [28–32], duration of active TB [33], and mortality rates [28–32], each of which is calibrated to account for parameter uncertainty (Table 1). PLWH are eligible for TPT regardless of LTBI infection status, reflecting guidance that TPT should not be delayed among PLWH for lack of available LTBI testing [34]. We assume that only people on ART initiate TPT.

**Table 1:** Dynamic transmission model input parameters. The input parameters used in the calibration with a mean value and range of 25% from the mean.

| Parameter description | Value [Range] | Reference(s) |
| --- | --- | --- |
| **Parameters that Impact Force of Infection with TB** |  |  |
| Number of effective contacts per infectious year for: |  | [36] |
| Males | 14 [10.5, 17.5] |  |
| Females | 14 [10.5, 17.5] |  |
| Relative transmissibility of TB (per year) for: |  | [26, 27] |
| HIV- | 1 (reference value) |  |
| HIV+, Not on ART, CD4 > 200 | 0.9 [0.675, 1.125] |  |
| HIV+, Not on ART, CD4 ≤ 200 | 0.6 [0.45, 0.75] |  |
| HIV+, On ART | 0.9 [0.675, 1.125] |  |
| Fraction of new TB infections that are MDR-TB | 0.037 [0.02775, 0.04625] | [37] |
| Diminished risk of acquiring latent DS-TB infection while on TPT for those who are uninfected | 0.43 [0.3225, 0.5375] | [38] |
| Increased risk of reinfection after developing active TB | 4 [3, 5] | [39] |
| Partially protective effect of LTBI against acquiring a new TB infection | 0.4 [0.3, 0.5] | [18, 26–28] |
| **Parameters that Describe TB Progression** |  |  |
| Duration of TPT course | 6 months (fixed value) | [18] |
| Duration of LTBI recently infected period | 2 years (fixed value) | [23] |
| Annual rate of progression from: |  |  |
| Recent LTBI to active TB | 0.025 [0.01875, 0.03125] | [43,44] |
| Remote LTBI to active TB | 0.001 [0.00075, 0.00125] | [45,46] |
| Relative risk of TB progression from LTBI to active TB: |  | [28–32] |
| HIV- | 1 (reference value) |  |
| HIV+, Not on ART, CD4 > 200 | 10 [7.5, 12.5] |  |
| HIV+, Not on ART, CD4 ≤ 200 | 17 [12.75, 21.75] |  |
| HIV+, On ART | 3 [2.25, 5.25] |  |
| Annual rates of progression among PLWH and on ART (IPT efficacy against DS TB) |  |  |
| LTBI on TPT to active TB | 0.0113 [0.0085, 0.0142] | [11] |
| LTBI after TPT to active TB | 0.0113 [0.0085, 0.0142] | [11] |
| Duration of active TB (if death does not occur) for: |  | [33] |
| HIV- | 2.0 years [1.5, 2.5] |  |
| HIV+, Not on ART, CD4 > 200 | 1.5 years [1.125, 1.875] |  |
| HIV+, Not on ART, CD4 ≤ 200 | 1.5 years [1.125, 1.875] |  |
| HIV+, On ART | 1.5 years [1.125, 1.875] |  |
| Rate of TB relapse per year | 0.01 [0.0075, 0.0125] | [47] |
| **Parameters that Describe HIV Progression** |  |  |
| HIV incidence time-varying rates<sup>†</sup> for: |  | [24] |
| Males | See appendix |  |
| Females | See appendix |  |
| Duration from HIV acquisition to CD4 <200: |  | [48] |
| Males | 7.72 years [5.79, 9.65] |  |
| Females | 10.25 years [7.6875, 12.8125] |  |
| **Parameters that Describe Mortality Rates** |  |  |
| Baseline mortality time-varying rates<sup>†</sup> for: |  | [24] |
| Males | See appendix |  |
| Females | See appendix |  |
| Increased risk of mortality for populations: |  | [30,49–54] |
| No active TB and HIV- | 1 (reference value) |  |
| Active TB, HIV- | 15.5 [11.625, 19.375] |  |
| Active TB, HIV+, Not on ART, CD4 > 200 | 26 [19.5, 32.5] |  |
| Active TB, HIV+, Not on ART, CD4 ≤ 200 | 50 [37.5, 62.5] |  |
| Active TB, HIV+, On ART | 18.5 [13.875, 23.125] |  |
| No active TB, HIV+, Not on ART, CD4 > 200 | 8 [6, 10] |  |
| No active TB, HIV+, Not on ART, CD4 ≤ 200 | 26 [19.5, 32.5] |  |
| No active TB, HIV+, On ART | 1.35 [1.2, 1.5] |  |
| **Parameters that Describe Entries and Exits due to Aging** |  |  |
| Proportion of the population that enters each compartment, time-varying | See appendix | [24] |

<sup>†</sup> Description of time-varying parameter calculations and mean values are in the appendix.

The population with active TB contributes to the force of infection for new LTBI among the susceptible population. The model allows different effective contact rates by gender, which are calibrated, but assumes homogeneous mixing in the population. The simulated population is kept constant at 100,000 by setting the total population entering the model equal to the total population exiting the model (due to aging out or dying). We ran the dynamic transmission model from the beginning of 1940 to the end of 2027, with a warmup and calibration period from 1940 through the end of 2017, and the intervention period from the start of 2018 through the end of 2027. We introduced HIV incidence in 1980.

We accounted for parameter uncertainty by calibrating 34 parameters using values 25% above and below the mean (Table 1). We used Latin hypercube sampling to generate 100,000 parameter sets for all 34 calibrated parameters [35]. We used each parameter set to execute the model from 1940-2017. Metrics from the model outputs for each parameter set were evaluated against 10 calibration target ranges in 2005 and 2017. The calibration target metrics were TB incidence and TB mortality by HIV status (HIV-positive and HIV-negative) and gender, and HIV prevalence by gender for adults aged 15-59 in KwaZulu-Natal, South Africa, from GBD 2019 [24]. We accepted parameter sets if all 20 metrics fell within the 95% uncertainty intervals of the calibration targets, resulting in 859 accepted parameter sets.

### Care Delivery Programs

We executed the model with the 859 accepted calibrated parameter sets for the three care delivery programs over a 10-year intervention period from the start of 2018 to the end of 2027. The programs differ in their gender-specific ART coverage and TPT initiation rates (Table 2). We assumed that programs took full effect at the start of the intervention period and were maintained over 10 years. Program 1 and Program 3 reflect the levels of ART coverage and TPT initiation rates observed in the facility-based care arm and the community-based care arm of the DO ART trial, respectively. Program 2 reflects the levels of ART coverage observed in Program 3, and assumes that TPT initation occurs at the facility-based care rate as observed in Program 1.

**Table 2:**
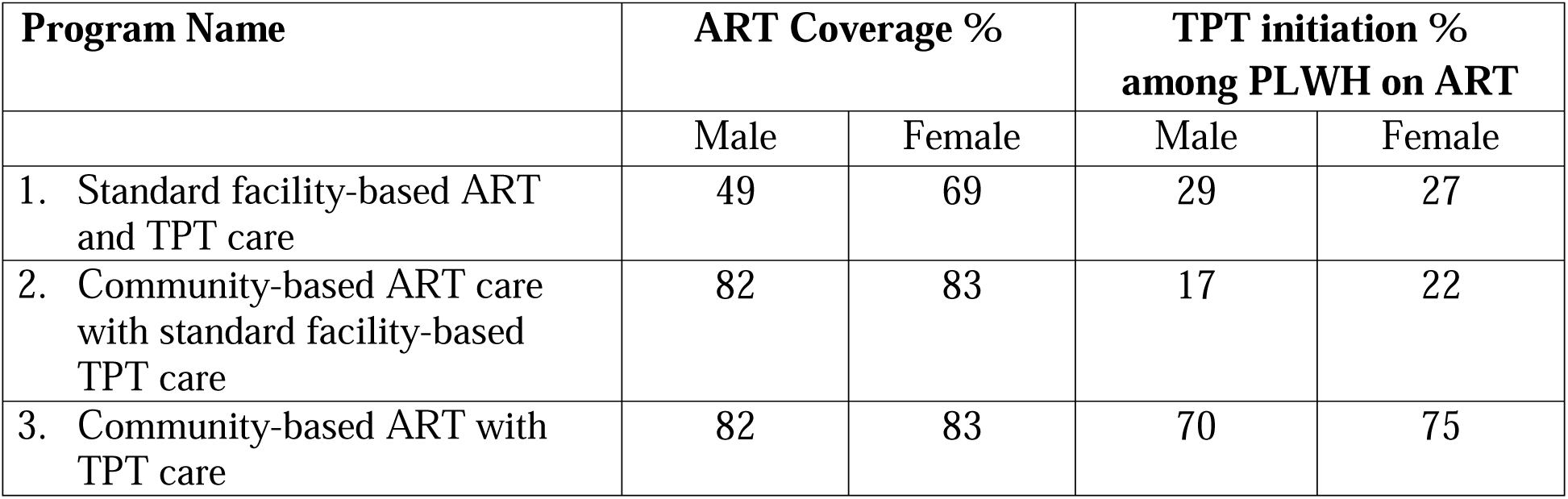
Facility-based and community-based care programs and parameters over the intervention period.

| Program Name | ART Coverage % |  | TPT initiation %<br>among PLWH on ART |  |
| --- | --- | --- | --- | --- |
|  | Male | Female | Male | Female |
| 1. Standard facility-based ART and TPT care | 49 | 69 | 29 | 27 |
| 2. Community-based ART care with standard facility-based TPT care | 82 | 83 | 17 | 22 |
| 3. Community-based ART with TPT care | 82 | 83 | 70 | 75 |

ART coverage estimates are based on the proportion of PLWH who achieve viral suppression under facility-based care (Program 1) and community-based care (Program 3) in the DO ART trial [18]. The proportion of PLWH on ART who achieve viral suppression and the proportion of PLWH diagnosed with HIV are based on population-based survey data [4]. The community-based care arm of the DO ART trial includes community-wide HIV testing, so we assume that all PLWH in Program 3 know their status. TPT initiation rates for Program 2 reflect the TPT initiation rates among all PLWH in Program 1, so that the number of individuals initiating TPT in Program 1 and Program 2 are comparable. Details are in the appendix.

### Cost Model

The cost model includes costs of TB and HIV-related care from the program perspective (Table 3). Further details of the component costs are in the appendix. The annual outpatient HIV care costs with community ART delivery is estimated at $310 per person based on a micro-costing study conducted during the DO ART trial under the “efficient at scale” scenario [18] and $249 per person for facility-based ART delivery. Inpatient and outpatient HIV care costs exclude the cost of care for TB, which is calculated separately [55]. The estimated HIV testing cost is based on the assumption that programs typically test multiple individuals for HIV before finding an eligible person to initiate ART screening. The cost of a six-month course of TPT includes the isoniazid cost [56], clinician time [57], the probability of developing drug-induced liver injury from isoniazid [11] and the costs of care for drug-induced liver injury [58]. TB care costs are estimated for standard treatment courses and reflect a higher treatment cost for MDR-TB [59] than for DS-TB [57]. We converted costs from the reported currency to 2018 US dollars using the consumer price index in South Africa and the United States related to the reported currency, and the currency conversion from the South African Reserve. We discounted future costs by 3%. We applied HIV- and TB-related costs to the populations in relevant model compartments at each time step and summed the total HIV and TB-related costs for each care program under the intervention period.

**Table 3:** Cost model input parameters. All values are presented as per person costs in 2018 USD and are rounded to the nearest dollar.

| Cost category | Value | Reference |
| --- | --- | --- |
| Annual outpatient HIV care costs for PLWH who are<sup>±</sup>: |  |  |
| Not on ART | 135 | [60] |
| On ART under facility-based care (Program 1) | 249 | [18] |
| On ART under community-based care (Programs 2 and 3) | 310 | [18] |
| Annual inpatient HIV care costs for PLWH who are<sup>±</sup>: |  |  |
| Not on ART, CD4 >200 | 62 | [55] |
| Not on ART, CD4 ≤ 200 | 162 | [55] |
| On ART (regardless of facility or community-based care) | 151 | [55] |
| Cost of HIV testing to find one person to initiate ART | 24 | [18, 55] |
| Cost of a 6-month course of TPT with isoniazid | 20 | [11,56–58] |
| Cost of a course of TB treatment for people with: |  |  |
| Drug-susceptible TB | 259 | [57] |
| Multidrug-resistant TB | 1889 | [59] |

<sup>±</sup>Inpatient and outpatient HIV care costs exclude the cost of care for TB, which is considered separately.

### Analysis

We calculated TB- and HIV-associated DALYs as the sum of years lived with disability (YLD) and years of life lost (YLL) during the 10-year intervention period using disability weights from GBD 2019 [61]. YLL is based on the gap between the time of death and the end of the intervention period. For example, if an individual died at the start of the eighth year of the intervention period, the YLL is two years. We discounted costs and DALYs accrued during the 10-year intervention period by 3%. The incremental cost-effectiveness ratios (ICERs) per TB death, per incident TB case, and per DALY averted are calculated for each care program over the intervention period. We assessed cost-effectiveness relative to the threshold of $590 USD per DALY averted based on the opportunity cost at the margin of the South African program [62, 63], as in Bershteyn, et al. [64]. We conducted one-way sensitivity analyses by varying the cost parameters in the model using the mean metrics from the 859 accepted parameter sets.

## Results

Figure 2 illustrates the projected annual TB incidence and mortality rates by gender from 1990 to 2027. In 2017, the year before the start of the intervention, the estimated annual incidence of active TB disease was slightly higher for women (1,406 per 100,000; range 1,141 – 1,577) than for men (1,165 per 100,000; range 989 – 1,362). In 2017, nearly 74.7% (1,050 out of 1,406) of women with incident TB were also living with HIV, compared to 62.4% (727 out of 1,165) of men with incident TB also living with HIV. However, the estimated TB mortality rate in 2017 was higher for men, with an estimate of 240 (range 174 – 309) per 100,000 men compared with 135 (range 103 – 188) per 100,000 women.

**Figure 2.**
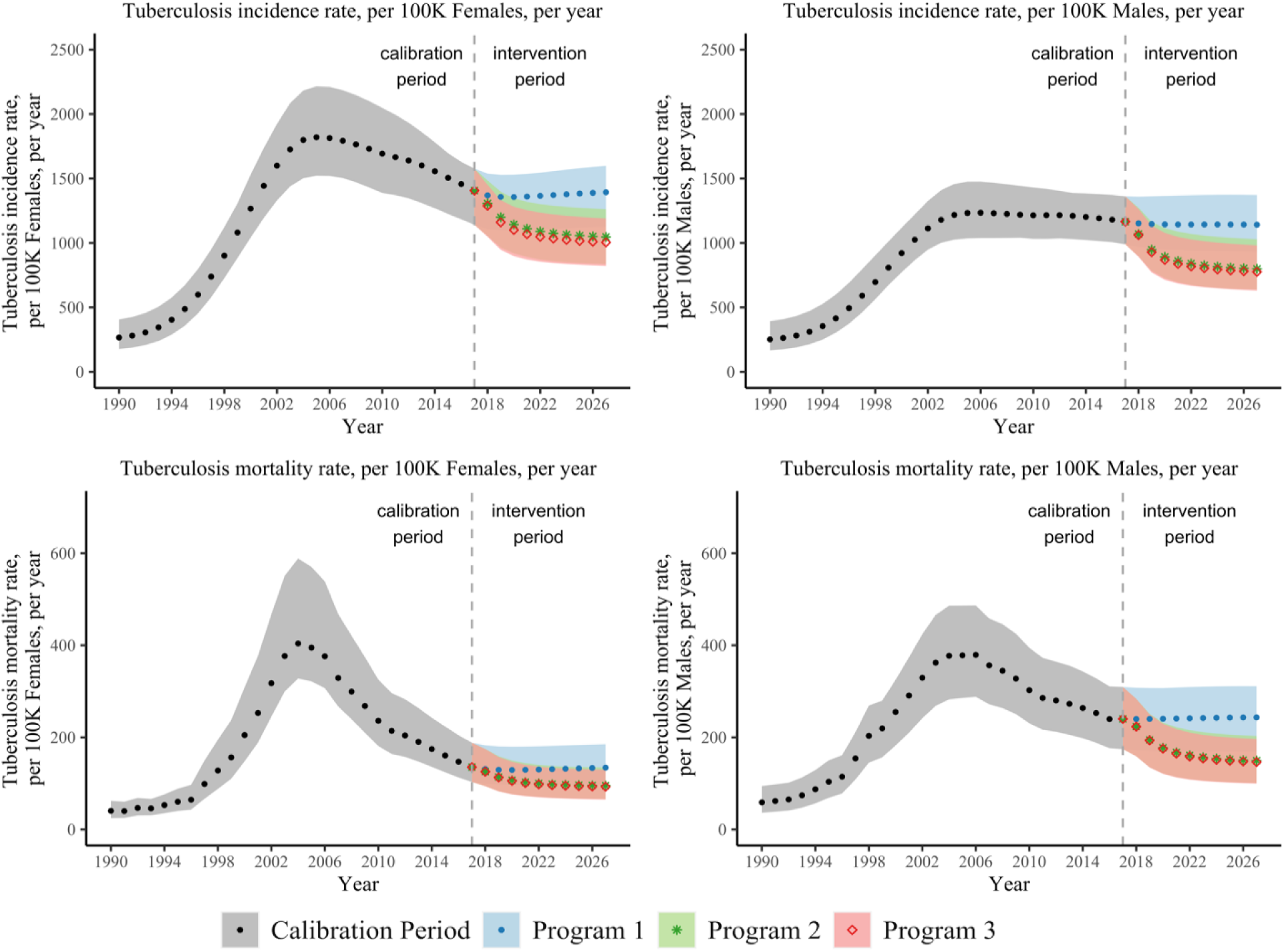
Estimated TB incidence and mortality by gender. The mean, maximum, and minimum yearly TB incidence and mortality rates from 1990-2017 over the 859 accepted parameter sets are shown in grey (dots). The mean, maximum, and minimum yearly TB incidence and mortality rates during the intervention period (2018-2027) over the 859 accepted parameter sets are illustrated by care-delivery program. During the intervention period, Program 1 (standard facility-based ART and TPT care) is shown in blue (dots), Program 2 (community-based ART care with standard facility-based TPT care) is shown in green (stars), and Program 3 (community-based ART with TPT care) is shown in red (diamonds).

Over the 10-year intervention period, an estimated 31,009 (range 25,674 – 39,021) PLWH received a course of TPT in Program 3 compared to 17,264 (range 14,156 – 21,255) in Program 1 and 18,221 (range 15,305 – 22,370) in Program 2 (see appendix for details). The estimated number needed to treat with TPT to prevent one case of active TB was 38.

As illustrated in Figure 2, the benefits of community-based care programs (Programs 2 and 3) compared to standard facility-based care (Program 1) are most apparent at the end of the intervention period. TB incidence and mortality rates by gender under each program in the last year of the intervention period are provided in Table 4. Community-based ART with TPT care (Program 3) reduced gender disparities in TB mortality compared to standard facility-based ART and TPT care (Program 1). TB mortality declined by 39.9% (range 32.2% – 46.3%) among men and 30.6% (range 25.3% - 36.5%) among women under Program 3 versus Program 1.

**Table 4:** Estimated TB incidence and mortality by care-delivery programs in the last year of the intervention period. The mean, maximum, and minimum yearly TB incidence and mortality rates are estimated over the 859 accepted parameter sets.

|  | **Program 1**<br>*Standard facility-based ART and TPT care* | **Program 2**<br>*Community-based ART care with standard facility-based TPT care* | **Program 3**<br>*Community-based ART with TPT care* |
| --- | --- | --- | --- |
|  | **Mean [Min, Max]** | **Mean [Min, Max]** | **Mean [Min, Max]** |
| **TB incidence** |  |  |  |
| TB incidence rate per 100,000 males | 1,141 [977, 1,301] | 799 [665, 950] | 775 [654, 902] |
| TB incidence rate per 100,000 females | 1,394 [1,185, 1,561] | 1,046 [883, 1,220] | 1,004 [848, 1,155] |
| **TB mortality** |  |  |  |
| TB mortality rate per 100,000 males | 243 [184, 300] | 149 [111, 191] | 146 [110, 184] |
| TB mortality rate per 100,000 females | 134 [103, 174] | 95 [70, 125] | 93 [69, 120] |

The model projected that community-based care programs also had indirect health benefits for people without HIV through reduced community TB transmission even though ART and TPT were only taken by PLWH. In 2027, TB incidence was 12.9% (range 8.3% – 18.1%) lower among men without HIV and 9.6% (range 4.7% – 15.1%) lower among women without in HIV under Program 3 versus Program 1. Estimated reductions in TB incidence and mortality for people without HIV by gender are detailed in the appendix.

In Figure 3, we illustrate the population-level impact of each program by comparing TB incidence and mortality rates, per 100,000 individuals over the intervention period. We estimated the impact of offering community-based ART (without changing TPT uptake) by comparing outcomes in Program 2 to Program 1. In 2027, Program 2 could reduce the TB incidence rate by 27.0% (range 21.3% – 34.1%) and the TB mortality rate by 34.6% (range 24.8% – 42.2%) compared to Program 1 (Figure 3). We estimated the impact of offering community-based TPT with ART by comparing outcomes in Program 3 to Program 2. In 2027, Program 3 could reduce TB incidence by an additional 3.6% (range 0.2% – 9.9%) and TB mortality rates by 2.2% (range 0.1% – 7.6%) compared to Program 2.

**Figure 3:**
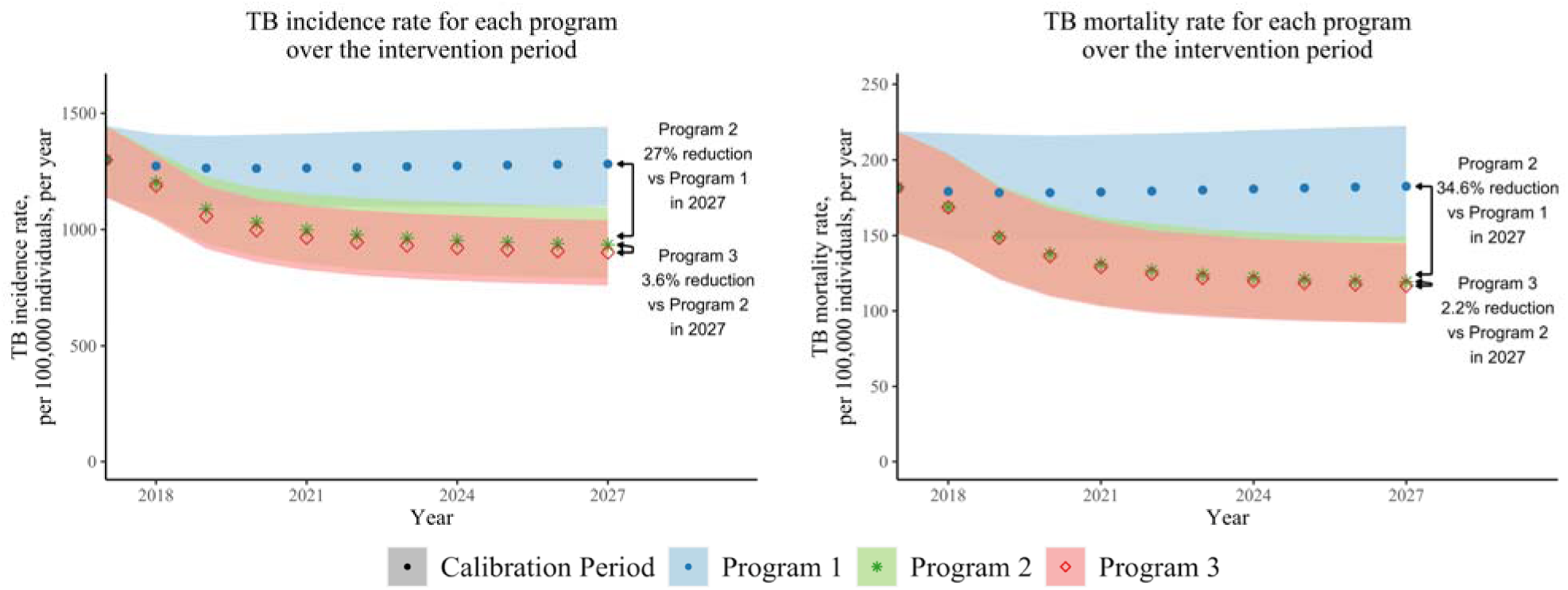
Estimated TB incidence and mortality by care-delivery program. The mean, maximum, and minimum yearly TB incidence and mortality rates are estimated over the 859 accepted parameter sets. Program 1 (standard facility-based ART and TPT care) is shown in blue (dots), Program 2 (community-based ART care with standard facility-based TPT care) is shown in green (stars), and Program 3 (community-based ART with TPT care) is shown in red (diamonds).

Health outcomes (DALYs, TB incident cases, TB deaths) and costs are summed over the 10-year intervention period for each care program, shown in Table 5. These program health outcomes and costs are used to calculate incremental health gains, costs, and ICERs for the outcomes of DALYs, incident TB cases, and TB deaths between facility-based and community-based care delivery programs as in Table 6.

**Table 5:** Cumulative health outcomes and costs over the intervention period by program. Values are the mean, minimum, and maximum values of the 859 accepted parameter sets. Health outcomes and costs are summed over the 10-year intervention period for a population of 100,000 individuals. Discounted values are presented in 2018 values and use an annual discount rate of 3%.

|  | **Program 1**<br>*Standard facility-based ART and TPT care* | **Program 2**<br>*Community-based ART care with standard facility-based TPT care* | **Program 3**<br>*Community-based ART with TPT care* |
| --- | --- | --- | --- |
|  | **Mean [Min, Max]** | **Mean [Min, Max]** | **Mean [Min, Max]** |
| **Health Outcomes and Costs (Undiscounted)** |  |  |  |
| DALYs (thousands) | 82.0 [70.5, 92.2] | 59.7 [52.4, 67.2] | 59.5 [52.2, 66.9] |
| TB incident cases (thousands) | 12.7 [10.5, 14.7] | 10.0 [8.1, 12.1] | 9.7 [8.1, 11.6] |
| TB deaths (thousands) | 1.8 [1.4, 2.3] | 1.3 [1.0, 1.7] | 1.3 [1.0, 1.7] |
| Program costs (millions in USD) | 99.3 [90.6, 114.1] | 118.2 [107.4, 135.5] | 118.4 [107.6, 135.8] |
| **Health Outcomes and Costs (Discounted)** |  |  |  |
| DALYs (thousands) | 73.4 [63.1, 82.5] | 53.8 [47.1, 60.5] | 53.6 [47, 60.2] |
| TB incident cases (thousands) | 11.2 [9.2, 12.9] | 8.8 [7.2, 10.7] | 8.6 [7.1, 10.3] |
| TB deaths (thousands) | 1.6 [1.2, 2] | 1.2 [0.9, 1.5] | 1.1 [0.9, 1.5] |
| Program costs (millions in USD) | 87.1 [79.5, 100.1] | 103.7 [94.3, 118.9] | 103.9 [94.4, 119.2] |

**Table 6:** Incremental health gains, costs, and incremental cost-effectiveness ratios (ICERs) between community-based and facility-based care delivery programs. Health outcomes and costs are summed over the 10-year intervention period for a population of 100,000 individuals. Values are the mean, minimum, and maximum values over 859 parameter sets. Discounted values are presented in 2018 values and use an annual discount rate of 3%.

|  | **Program 2 vs.**<br>**Program 1**<br>*Community-based ART with standard facility-based TPT care vs. facility-based ART and TPT care* | **Program 3 vs.**<br>**Program 1**<br>*Community-based ART with TPT care vs. facility-based ART and TPT care* | **Program 3 vs.**<br>**Program 2**<br>*Community-based ART with TPT care vs. community-based ART with facility-based TPT care* |
| --- | --- | --- | --- |
|  | **Mean [Min, Max]** | **Mean [Min, Max]** | **Mean [Min, Max]** |
| **Incremental Health Gains and Costs (Undiscounted)** |  |  |  |
| DALYs averted (thousands) | 22.3 [18.1, 27] | 22.5 [18.2, 27.3] | 0.2 [0, 0.5] |
| TB incident cases averted (thousands) | 2.7 [2.1, 3.5] | 3.0 [2.3, 3.7] | 0.3 [0, 0.9] |
| TB deaths averted (thousands) | 0.5 [0.3, 0.7] | 0.5 [0.4, 0.7] | 0.0 [0.0, 0.1] |
| Incremental costs (millions in USD) | 18.8 [16.8, 21.5] | 19.0 [17.0, 21.7] | 0.2 [0.1, 0.3] |
| **Incremental Health Gains and Costs (Discounted)** |  |  |  |
| DALYs averted (thousands) | 19.7 [15.9, 23.8] | 19.8 [16.0, 24.0] | 0.1 [0.0, 0.4] |
| TB incident cases averted (thousands) | 2.3 [1.8, 3.0] | 2.6 [2.0, 3.2] | 0.3 [0.0, 0.8] |
| TB deaths averted (thousands) | 0.4 [0.3, 0.6] | 0.4 [0.3, 0.6] | 0.0 [0.0, 0.1] |
| Incremental costs (millions in USD) | 16.6 [14.8, 18.9] | 16.7 [14.9, 19.1] | 0.2 [0.1, 0.3] |
| **Incremental Cost-effectiveness Ratios (Health gains and costs are discounted by 3%)** |  |  |  |
| Cost per DALY averted (in USD) | 843<br>[706, 1,017] | 846<br>[709, 1,012] | 1,967<br>[244, 20,282] |
| Cost per incident TB case averted (in USD) | 7,192<br>[5,161, 10,010] | 6,498<br>[4,823, 9,304] | 1,109<br>[107, 13,012] |
| Cost per TB death averted (in USD) | 40,692<br>[26,116, 62,157] | 39,373<br>[25,606, 57,020] | 19,737<br>[1,783, 470,597] |

The community-based ART with TPT care program (Program 3) did not meet the cost-effectiveness threshold of $590 USD per DALY averted with the input parameter costs given in Table 3. Figure 4 shows the impact of varying costs on the discounted incremental cost per DALY averted under Program 3 versus Program 1. The discounted incremental cost per DALY was most sensitive to the cost of outpatient HIV care. If the cost of annual outpatient care for PLWH on ART under community-based ART programs (Programs 2 and 3) were to decrease from $310 to $283, or if the cost of annual outpatient care for PLWH on ART under standard facility-based ART and TPT care (Program 1) were to increase from $249 to $260, then the discounted incremental cost per DALY would achieve the cost-effectiveness threshold.

**Figure 4:**
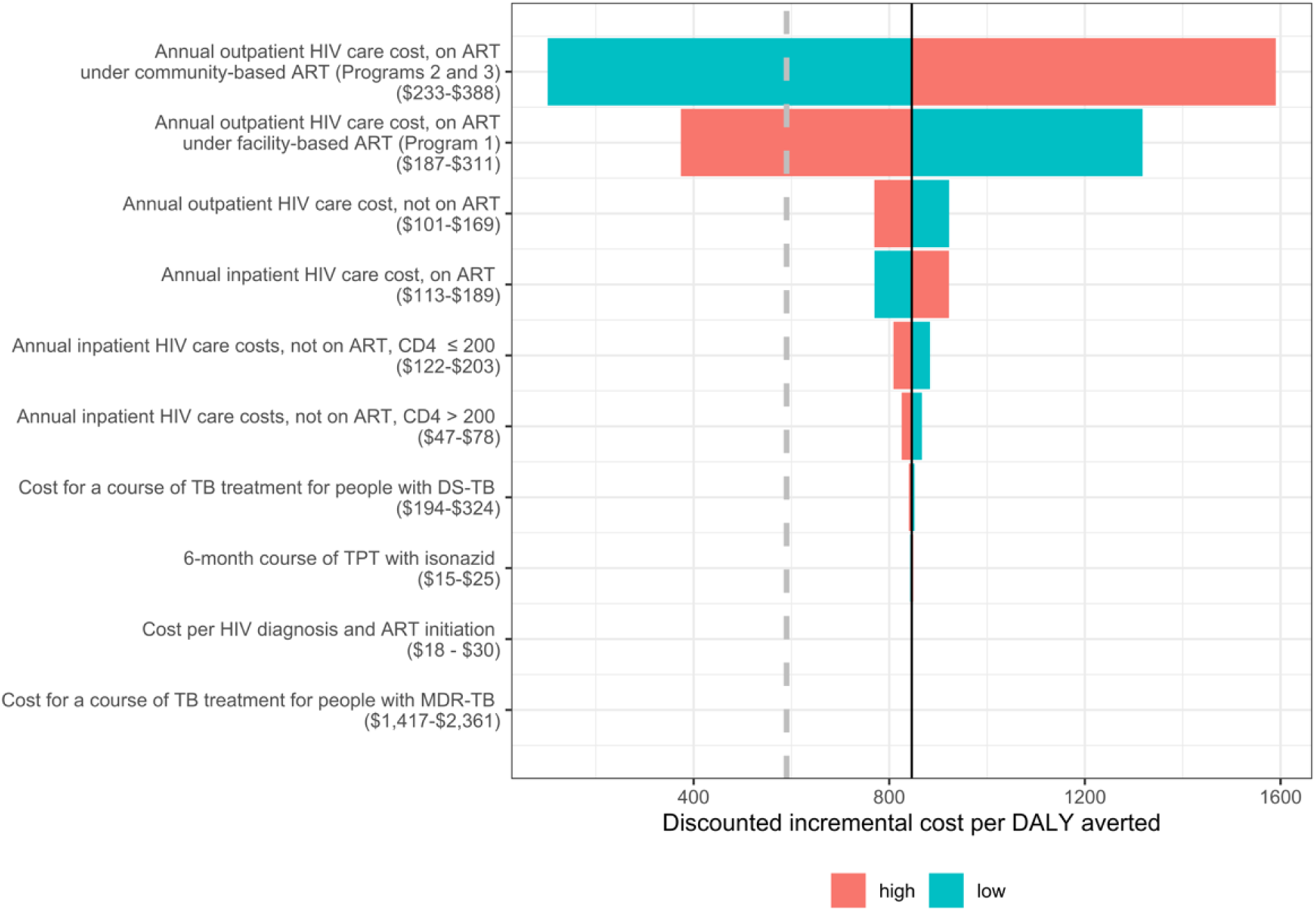
Sensitivity analysis of the discounted incremental cost per DALY averted by community-based ART with TPT care (Program 3) versus standard facility-based ART and TPT care (Program 1) over the intervention period. The solid line represents the mean discounted incremental cost per DALY averted over the 859 accepted parameter sets of $846 USD per DALY averted. The dashed vertical line represents the cost-effectiveness threshold of $590 USD per DALY averted. The horizontal bars represent the discounted incremental cost per DALY averted at bounds of 25% above of the modeled cost parameter (high) and 25% below the modeled cost parameter (low). All costs are in 2018 USD.

## Discussion

Scaling up community-based ART with TPT care to the population level in a high HIV-TB burden setting such as KwaZulu-Natal, South Africa, could avert substantial HIV and TB morbidity and mortality, if equivalent effectiveness is maintained as demonstrated in the DO ART trial. The increase in ART coverage achieved through community-based care reduced cumulative TB disease by 21% over ten years, and community-based TPT care reduced it further by 3%. Additionally, though the interventions were only directly delivered to PLWH, the model captured indirect benefits of reduced TB incidence disease and mortality among people without HIV under the community ART and TPT scenarios. Prior modeling analyses in South Africa have also found that ART and TPT scale-up would be projected to reduce TB incidence among people living with and without HIV [65].

Gender disparities in HIV and TB outcomes in this setting are complex and are reflected in the model results. Specifically, the higher HIV prevalence among women (39%) compared to men (20%) in our model resulted in higher incidence and mortality of HIV-associated TB among women than among men, while HIV-negative women had lower TB incidence, prevalence, and mortality than HIV-negative men. The larger increase in ART coverage under the community-based care programs among men resulted in larger percent reductions in 10-year TB-associated mortality among men than among women compared to standard clinic-based care. Gender-specific model parameters included effective contact rates, HIV prevalence, HIV disease progression, ART coverage, and baseline (non-HIV and non-TB associated) mortality, though other features of TB epidemiology may also differ by gender, including care seeking behavior that may impact active TB duration and case fatality ratios. A recent gender-specific TB model in a setting with a low HIV prevalence found that interventions targeted to risk factors with a higher prevalence among men (e.g. tobacco smoking and harmful alcohol use) achieved the greatest projected TB incidence reductions among men, but also substantially reduced TB incidence among women and children [66].

Our cost analysis found that community-based ART with TPT care was cost ineffective relative to the cost-effectiveness threshold based on the opportunity cost at the margin of the South African programme [64]. The sensitivity analysis of cost parameters indicated that variation in the costs of outpatient HIV care for PLWH on ART substantially affected the ICER. If the annual per person costs of outpatient HIV care for PLWH on ART under community-based care were to decrease from $310 to $283 or the annual per person costs of outpatient HIV care for PLWH on ART under facility-based care were to increase to from $249 to $260, then community-based ART with TPT (Program 3) could be cost effective. Other community-based interventions, including interval TB/HIV screening and linkage to care, were found to be cost-effective in a modeling analysis set in South Africa, though these did not include community-based HIV treatment and were evaluated against a higher GDP-based cost-effectiveness threshold [68].

Strengths of this analysis include that it is based in findings from the recent DO ART randomized controlled trial conducted in this setting. Additionally, the model is calibrated to gender- and province-specific estimates of TB and TB-HIV incidence and mortality at two time points that capture the declines in HIV-associated TB incidence and mortality in the setting of increased ART coverage [5]. To reflect these dynamics, we included time varying parameters for HIV incidence, prevalence, and ART coverage that incorporate historical changes in ART availability. Additionally, the model incorporates uncertainty in model parameters. Uncertainty remains in key aspects of TB epidemiology, including the rate of progression from latent to active TB, which is represented differently across models [69]. Finally, by using a model that differentiates compartments by gender, we were able to stratify outcomes by gender, which is important for evaluating health program impacts on gender equity in health outcomes.

This analysis also has limitations. First, we assumed that the modeled care programs could take effect immediately and be sustained over ten years, which may not be feasible for already-strained health care workforce and budgets. However, dramatic increases in TPT initiation and completion have been observed in some HIV high-incidence settings through focused outreach campaigns [70]. Second, the model does not differentiate groups by age despite differences in HIV and TB incidence, prevalence, and mortality by age group due to the complexity of calibration across this additional dimension. Third, for simplicity, we assumed that people taking TPT would not be protected from infection with an MDR-TB strain, though some evidence suggests that they may have partial protection from a resistant strain [71]. Finally, the economic analysis is from a health system perspective that does not include patient costs, despite numerous studies documenting patient costs and avoiding catastrophic costs as a key focus of TB goals [72].

## Conclusions

If community-based ART with TPT care was scaled up to achieve similar levels of ART and TPT coverage as in the DO ART trial, the TB and HIV dynamic transmission model predicts that TB disease would be reduced by 24% and deaths reduced by 28% over ten years in KwaZulu-Natal, South Africa. Differentiated models of HIV care can substantially reduce TB morbidity and mortality and reduce gender disparities in TB.

## Authors’ contributions

JMR and RVB conceptualized the study. CG formulated and programmed the model, with guidance from ZBZ and parameterization from JMR, CG, CJB, DWR, AES, and DWD. AvH, DAR, AES, and RVB provided critical interpretation of the DO ART trial data. All authors contributed to scenario definition and analysis planning. JMR wrote the first draft of the manuscript. CG wrote the first draft of the appendix. All authors contributed to the interpretation of the findings, provided critical review and revisions, and approved the final version.

## Data Availability

All data produced in the present study are available upon reasonable request to the authors.

## Additional files

Additional file 1: Appendix

The appendix file contains additional details about the study methods and additional results.

