## Supplementary appendix for "Preventing tuberculosis with community-based care in an HIV-endemic setting: a modeling analysis"

May 5, 2023

**2 Dynamic Transmission Model Equations 10**

### 1 Model Description

#### 1.1 Dynamic Transmission Model

The model considers eight tuberculosis (TB) compartments, two TB drug resistance (DR) compartments, four human immunodeficiency virus (HIV) compartments, and two gender compartments described in Section 1.1.1. The model considers a population of 100,000 adults ages 15-59 in KwaZulu-Natal, South Africa, as described in Section 1.1.2. Rates of flow between each compartment that reflect TB and HIV disease progression and entries and exits from the model are summarised in Section 1.1.3. Outputs generated by the model are described in Section 1.1.4.

##### 1.1.1 Model Compartments

*Set of Tuberculosis (TB) Compartments*: $t \in TB$

The model consists of uninfected, latent TB infection (LTBI), active TB, and recovered/treated TB compartments. We separate LTBI compartments into recent (less than two years since exposure) and remote (at least two years after exposure to TB) to account for the difference in risk of progression [12, 15, 17, 29]. Those who are uninfected or have LTBI can initiate TB preventative therapy (TPT). We include a compartment for LTBI after TPT to account for the reduced risk of progression to active TB after completion of TPT [3, 13, 38]. This yields eight TB compartments. We define the set of TB compartments as:

1. Uninfected, not on TPT
2. Uninfected, on TPT
3. LTBI, infected recently (exposed within the past two years)
4. LTBI, infected remotely (exposed more than two years ago)
5. LTBI, on TPT
6. Active TB
7. Recovered/Treated
8. LTBI, after TPT

*Set of Tuberculosis Drug Resistance (DR) Compartments*: $r \in DR$

The model differentiates drug-susceptible TB (DS-TB) and multidrug-resistant TB (MDR-TB) infections. We define MDR-TB as resistant to TPT with isonazid and rifampicin. Uninfected individuals on TPT have a reduced risk of acquiring DS LTBI, but we assume no reduced risk of acquiring MDR LTBI [14]. Individuals with DS LTBI on TPT have a reduced risk of progression to active TB, but we assume those with MDR LTBI on TPT do not benefit from a reduced risk of progression to active TB [2, 6, 9, 26, 45, 49]. We define the set of DR compartments as:

1. Drug-susceptible (DS)
2. Multidrug-resistant (MDR)

For individuals who are uninfected (TB compartments 1 or 2), it is not possible to distinguish by DR status, so all individuals in TB compartments 1 and 2 are assigned to DR compartment 1. We only allow individuals with DS LTBI (TB compartments 3 or 4 and DR compartment 1) to enter LTBI on TPT (TB compartment 5) and subsequently LTBI after TPT (TB compartment 8). As such, the possible combinations of TB and DR compartments are:

TB compartment 1, DR compartment $r = 1$ only

TB compartment 2, DR compartment $r = 1$ only
TB compartment 3, both DR compartments $r = 1$ and $r = 2$
TB compartment 4, both DR compartments $r = 1$ and $r = 2$
TB compartment 5, DR compartment $r = 1$ only
TB compartment 6, both DR compartments $r = 1$ and $r = 2$
TB compartment 7, both DR compartments $r = 1$ and $r = 2$
TB compartment 8, DR compartment $r = 1$ only

*Set of HIV Compartments*: $h \in HIV$

The model consists of four HIV compartments that are differentiated by their relative transmissibility of TB, risk of progression from LTBI to active TB, duration of active TB and mortality rates [2, 6, 9, 21, 26, 30, 34–36, 45, 49]. Only people living with HIV (PLWH) are eligible to initiate ART, and only those on ART initiate TPT treatment [7]. We define the set of HIV compartments as:

1. HIV-
2. HIV+, not on ART, CD4 $> 200$
3. HIV+, not on ART, CD4 $\leq 200$
4. HIV+, and on ART

*Set of Gender Compartments*: $g \in G$

The model consists of two gender compartments that are differentiated by effective contact rates, rate of TPT and ART initiation, HIV incidence, CD4 decline, and mortality rates [19, 20, 25, 28, 39, 40]. We define the set of gender compartments as:

1. Male
2. Female

##### 1.1.2 Population

The model includes an adult population between the ages of 15 and 59 with no immigration or emigration. We define $N_{t,r,h,g}(\tau)$ as the total population in TB compartment $t$, DR compartment $r$, HIV compartment $h$, and gender compartment $g$ at time $\tau$. For the purpose of this model, we keep the size of the population constant at all time steps such that

$$\sum_{t \in TB} \sum_{r \in DR} \sum_{h \in HIV} \sum_{g \in G} N_{t,r,h,g}(\tau) = 100,000.$$

We keep the population constant by setting the total amount of population entering the model to the total amount exiting the model (due to aging out or dying). Some proportion of the population aging into the model ages into TB compartments 1 (Uninfected, not on TPT), 3 (LTBI, infected recently), 4 (LTBI, infected remotely), and 6 (active TB) according to yearly population, LTBI prevalence, and TB prevalence estimates by gender. LTBI prevalence and TB prevalence estimates are differentiated by drug-resistance when indicated by estimates. Some proportion of the population ages into HIV compartments 1 (HIV-) and 2 (HIV+, not on ART, CD4>200) according to yearly HIV prevalence estimates. We account for changes in population characteristics of those aging into the model over time by adjusting the proportion of the population aging into each TB,

HIV, and gender compartment according to yearly estimates and dynamic parameters (e.g., HIV incidence rates) from the 2019 Global Burden of Disease study (GBD 2019) for KwaZulu-Natal, SA [4].

###### 1.1.3 Model Transitions

Rates of flow between each compartment are governed by differential equations as described in Section 2.2. Transitions that describe TB and HIV disease progression are illustrated in Figure 1 and Figure 2. Although not depicted in Figure 1 and Figure 2, there are entries to reflect those aging into and exiting the model as described at the end of this section. The description and notation for the parameters used in these transitions are summarized in Table 1, where time-varying parameters are indicated by $\tau$. The time-varying parameters in Table 1 that are calculated using population states, specifically TB force of infection rates for populations in DR compartment $r$ at time $\tau$, $\lambda_r(\tau)$, ART initiation rates for gender $g$, $\eta_{2,4,g}(\tau)$ and $\eta_{3,4,g}(\tau)$, the total population entering the model, $B(\tau)$, and rates of exits from mortality and aging out, $\alpha^{out}_{t,r,h,g}(\tau)$, are calculated using the equations defined in Section 2.1. The parameters that are directly impacted by care programs (ART and TPT initiation rates) are highlighted in green in Figure 1, Figure 2, Table 1, and Table 2. The equations used to describe model transitions are described in Section 2.2.

*TB disease progression*

Figure 1 illustrates transitions to and from each TB compartment with words and mathematical notation as in Table 1. TPT initiations are highlighted in green to emphasize that care programs directly impact them. The primary focus of the analysis is the impact of different programs on active TB (TB compartment 6), as highlighted in red, including TB incidence rates (the number of individuals moving into the active TB compartment per year) and TB mortality rates (the number of individuals who die in the active TB compartment, per year).

Individuals exit TB compartment 1 (uninfected, not on TPT) through TB infection or TPT initiation. While all individuals in TB compartment 1 are arbitrarily assigned to DR compartment 1 ($r = 1$), when transitioning to TB compartment 3 (LTBI, infected recently), the force of infection is differentiated by drug resistance status ($\lambda_r(\tau)$), which is based on a proportion of those infected with DS-TB and MDR-TB. Uninfected individuals who initiate TPT enter TB compartment 2 (uninfected, on TPT) and DR compartment 1 ($r = 1$). Individuals in TB compartment 2 (uninfected, on TPT) that contract TB while on TPT and transition to TB compartment 3 (LTBI, infected recently) are differentiated by drug resistance status according to the force of infection ($\lambda_r(\tau)$) and a parameter that accounts for the reduced risk of infection of DS-TB while on TPT ($\iota_r$).

The transition rate for all those that move into TB compartment 3 (LTBI, infected recently) by reinfection (from TB compartments 4, 7, and 8) accounts for the individual's most recent TB strain type. The transition rate of reinfection from TB compartments 4 and 8 is determined by the force of infection $\lambda_r(\tau)$ and the partially-protective effect of LTBI against acquiring a new TB infection ($\zeta$). However, individuals in TB compartment 7 (Recovered/Treated) have an increased risk of reinfection, where the force of infection is modified by an increased risk of reinfection ($\xi$).

Individuals in TB compartment 3 (LTBI, infected recently) will on average, progress to TB compartment 4 (LTBI, infected remotely) after two years. Some proportion of individuals in TB compartment 3 will progress to TB compartment 6 (active TB) or TB compartment 5 (LTBI, on TPT). We assume only PLWH on ART will initiate TPT, which is captured by the TPT initiation rates. We assume individuals with MDR LTBI do not benefit from a reduced risk of progression to active TB (TB compartment 6) by disallowing those with MDR LTBI to move into TB compartment

| Notation | Description |
|---|---|
| **Parameters that impact TB infection** | |
| $\lambda_r(\tau)$ | The TB force of infection for populations in DR compartment $r$ at time $\tau$ |
| $\iota_r$ | Diminished risk of acquiring a latent TB infection for those exposed to DS-TB while on TPT, such that $0 < \iota_1 < 1$ and $\iota_2 = 1$. |
| $\xi$ | Increased risk of reinfection after recovery/treatment of active TB |
| $\zeta$ | Partially-protective effect of LTBI against acquiring a new TB infection |
| **Parameters that describe TB progression** | |
| $\omega$ | Rate of moving off of TPT, per year |
| $\gamma_r$ | Indicator to reflect that those infected with MDR LTBI do not yield benefits of TPT, such that $\gamma_1 = 1, \gamma_2 = 0$ |
| $\pi_{3,4}$ | Rate of TB progression from recent LTBI to remote LTBI, per year |
| $\pi_{3,6}$ | Rate of TB progression from recent LTBI to active TB, per year |
| $\pi_{4,6}$ | Rate of TB progression from remote LTBI to active TB, per year |
| $\theta_h$ | Relative risk for TB progression from LTBI to active TB by HIV compartment $h$, where those who are HIV negative ($h = 1$) progress at the base rate, $\theta_1 = 1$, and $\theta_h$ for $h = 2, 3, 4$ accounts for an increased risk of TB progression, $\theta_h > 1$ |
| $\pi_{5,6}$ | Rate of TB progression from LTBI on TPT to active TB, per year |
| $\pi_{7,6}$ | Rate of relapse from recovered/treated to active TB, per year |
| $\pi_{8,6}$ | Rate of TB progression from LTBI after TPT to active TB, per year |
| $\pi_{6,7}$ | Rate of recovery/treatment from active TB, per year |
| $\upsilon_h$ | Relative reduction of recovery/treatment rate by HIV compartment $h$, to reflect delays in TB treatment, $0 < \upsilon_h \leq 1$ |
| $\kappa_{h,g}(\tau)$ | Rate of TPT initiation from HIV compartment $h$ for gender $g$ at time $\tau$, per year, where only individuals in HIV compartment 4 (HIV+, on ART) initiate TPT, such that $\kappa_{h,g} = 0$ for $h = \{1, 2, 3\}$, and for all $g$ |
| **Parameters that describe HIV progression** | |
| $\eta_{1,2,g}(\tau)$ | HIV incidence rate for gender $g$ at time $\tau$, per year, HIV compartment 1 (HIV-) to HIV compartment 2 (HIV+, not on ART, CD4 > 200) |
| $\eta_{2,3,g}$ | Rate of HIV progression from HIV compartment 2 (HIV+, not on ART, CD4 > 200 to HIV compartment 3 (HIV+, not on ART, CD4 $\leq$ 200 for gender $g$ |
| $\eta_{2,4,g}(\tau)$ | ART initiation rate from HIV compartment 2 (HIV+, not on ART, CD4 > 200) to HIV compartment 4 (HIV+, on ART) for gender $g$ at time $\tau$, per year. |
| $\eta_{3,4,g}(\tau)$ | ART initiation rate from HIV compartment 3 (HIV+, not on ART, CD4 $\leq$ 200) to HIV compartment 4 (HIV+, on ART) for gender $g$ at time $\tau$, per year. |
| **Parameters that describe entries and exits from the population** | |
| $B(\tau)$ | Population aging into the model at time $\tau$ |
| $\alpha^{out}_{t,h,g}(\tau)$ | Rate of the population exiting the model due to mortality or aging out in TB compartment $t$, HIV compartment $h$, and gender compartment $g$ at time $\tau$, per year. |
| $\alpha^{in}_{t,r,h,g}(\tau)$ | Proportion of population that enters into TB compartment $t$, DR compartment $r$, HIV compartment $h$, and gender compartment $g$ at time $\tau$ due to aging in. |

Table 1: Description and notation for parameters used in the equations that describe TB and HIV disease progression, treatment rates, and entries and exits from the model. Time-varying parameters are notated as functions of $\tau$. Parameters that depend on the care program are highlighted in green. TB force of infection and HIV incidence rates are highlighted in blue to indicate the care program indirectly impacts them.

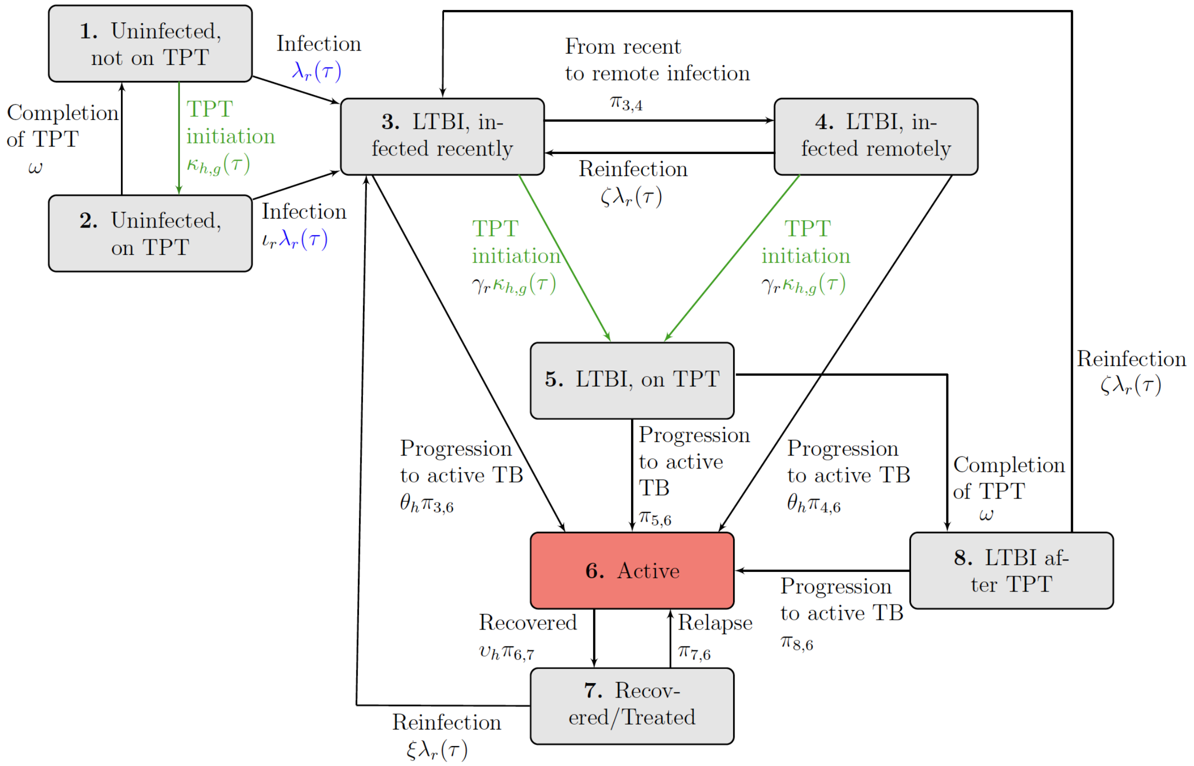

Figure 1: Differential equations govern rates of flow between each compartment. Although not visualized here, each tuberculosis (TB) compartment is stratified across two TB drug-resistance compartments, four human immunodeficiency virus (HIV) compartments, and two genders. The latent TB infection (LTBI) compartment is distinguished by those infected within two years, considered recent, and more than two years, considered remote. Individuals can age into TB compartments 1 (uninfected, not on TPT), 3 and 4 (LTBI, recent and remote), and 6 (active TB), and HIV compartments 1 (HIV-) and 2 (HIV+, not on ART, CD4 > 200). Individuals can age out or die from any compartment. TB preventative therapy (TPT) initiations are highlighted in green to emphasize that care programs directly impact them. TB force of infection is highlighted in blue to indicate the care program indirectly impacts them. The active TB compartment is highlighted in red to emphasize the compartment capturing TB incident cases and TB mortality. TB incidence rates are based on the number of individuals transitioning into the active TB compartment. TB mortality rates are based on the number of individuals in the active TB compartment that die.

5 (LTBI, on TPT) and subsequently TB compartment 8 (LTBI, after TPT). Those with LTBI who complete their course of TPT move into TB compartment 8 (LTBI after TPT) to account for the reduced risk of progression even after completing their TPT course.

Individuals who progress to TB compartment 4 (LTBI, infected remotely) have a reduced risk of progression to active TB compared to those in TB compartment 3 (LTBI, infected recently). PLWH have an increased risk of progression to TB compartment 6 (active TB). Rates of recovery are differentiated by HIV status to represent varying expected delays to treatment. A proportion of those who recover from active TB will relapse or become reinfected.

*HIV disease progression*

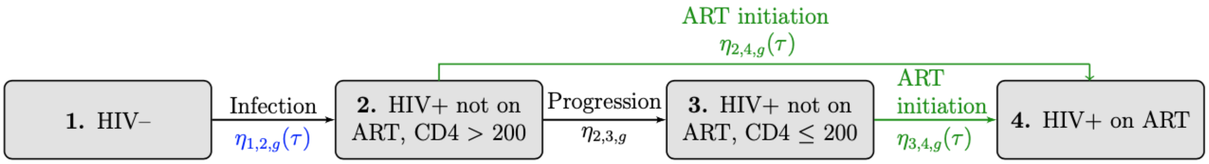

Figure 2: Illustration of human immunodeficiency virus (HIV) transitions between HIV compartments for each gender compartment $g$ at time $\tau$. Antiretroviral therapy (ART) initiation rates (highlighted in green) are directly impacted by ART coverage for each care program. ART coverage is used to calculate ART initiation rates so that the proportion of PLWH on ART by gender corresponds to ART coverage assumptions. HIV incidence rates are highlighted in blue to indicate the care program indirectly impacts them. Although not visualized here, each HIV compartment is stratified across eight TB, two TB drug resistance, four HIV, and two gender compartments.

Figure 2 illustrates transitions to and from each HIV compartment in words and with mathematical notation as defined in Table 1. ART initiation rates are highlighted in green to indicate that the care program directly impacts them. Those in HIV compartment 1 (HIV-) acquire HIV according to incidence rate estimates over time for each gender. Some proportion of those in HIV compartment 2 (HIV+, not on ART, CD4 $>$ 200) will initiate ART and transition into HIV compartment 4 (HIV+, on ART), and others experience CD4 decline and transition into HIV compartment 3 (HIV+, not on ART, CD4 $\leq$ 200). Net ART initiations from HIV compartment 2 (HIV+, not on ART, CD4 $>$ 200) and HIV compartment 3 (HIV+, not on ART, CD4 $\leq$ 200) are based on ART coverage estimates over time by gender, and the guidance on eligibility for PLWH to initiate ART based on their CD4 count as outlined in Section 2.1.2.

*Entries and exits*

Although not depicted in Figure 1 and Figure 2, transitions for entries and exits to the model are used to maintain constant population size. We keep the size of the population constant by setting entries from aging-in equal to exits from aging out or dying. Individuals can age into the model into TB compartments 1 (Uninfected, not on TPT), 3 (LTBI, infected recently), 4 (LTBI, infected remotely), and 6 (active TB). Also, individuals can age into HIV compartments 1 (HIV-) and 2 (HIV+, not on ART, CD4 $>$ 200). The proportion of the population aging into TB and HIV compartments by DR compartment (for those aging into TB compartments 3, 4, and 6) and gender is accounted for based on estimates from GBD 2019 for KwaZulu-Natal, South Africa [4]. All compartments include exit transitions from aging out or dying.

###### 1.1.4 Model Outputs

The model projects TB incidence, TB mortality, TB and HIV prevalence, and disability-adjusted life years (DALYs) based on population states and transition rates. DALYs represent the years of life lost (YLL) and years lived with disability (YLD) for TB and HIV disease. The equations used to calculate these metrics are described in Section 4. During the intervention period, we generate model outputs for three care programs described in the next section, Section 1.2.

##### 1.2 Care Programs

We executed the model for three care programs, including:

- *Program 1:* Standard facility-based ART and TPT care
- *Program 2:* Community-based ART care with standard facility-based TPT care, and
- *Program 3:* Community-based ART with TPT care.

The parameters that depend on the care program are TPT initiation rates and ART coverage. These parameters are estimated using data from the DO ART trial [7]. Community-based care programs aim to increase ART coverage and TPT initiation. The DO ART trial participants were recruited through HIV testing at clinics, community locations, and at home. While all PLWH in the DO ART trial were diagnosed with HIV, in our model we include PLWH who do not know their status. ART coverage is estimated based the HIV viral load of participants. We assume some PLWH on ART do not achieve viral suppression. Only PLWH not on ART (in HIV compartments 2 and 3) initiate ART, and only PLWH on ART (in HIV compartment 4) initiate TPT.

We evaluate these programs over a 10-year intervention period. We assume that programs take full effect at the start of the 10-year intervention period, such that ART coverage and TPT initiation assumptions for each program immediately increase at the beginning of the intervention period and are held constant over the 10-year intervention period. The standard of care, Program 1, reflects the levels of ART coverage and TPT initiation observed among participants in the standard facility-based care arm of the DO ART trial [7]. Community-based ART was tested in the DO ART trial with a community-based TPT care program (Program 3); however, we additionally test community-based ART without community-based TPT (Program 2) by setting TPT initiation rates among all PLWH to the same rate as the TPT standard of care scenario, so that the number of individuals initiating TPT in Program 1 and Program 2 are comparable. Additional detail about how the DO ART data are used to parameterize the modeled care programs is provided in Section 5.3. In Figure 1, Figure 2, and Table 1, parameters that are directly impacted by the care program are highlighted in green. ART initiation rates are calculated based on ART coverage using the equations described in Section 2.1.

##### 1.3 Cost Model

The cost model generates costs of TB and HIV care, including the costs to administer ART and TPT and HIV testing costs. The cost to distribute ART is based on the number of individuals in HIV compartment 4 (HIV+, on ART) and the yearly cost to provide ART under community-based and facility-based care programs. Costs for community-based ART care are higher than costs for standard facility-based ART care to reflect additional costs for the additional cadre of health workers and other resources needed to deliver community-based care. The annual cost of community ART care is based on a micro-costing study conducted during the DO ART trial under the "efficient at scale" scenario [7].

The cost to administer TPT is based on the number of individuals in TB compartment 2 (uninfected, on TPT) and TB compartment 5 (LTBI, on TPT) and the medication costs, provider time for counseling, and provider laboratory costs associated with drug-induced liver injury from TPT over a six-month course of TPT. Since the TPT community-based care program is nested within the ART community-based care program, costs to administer TPT through community-based care are the same as facility-based care.

Costs for TB care are defined for a course of TB treatment. They are differentiated by DS-TB and MDR-TB infections to account for differentiated treatment regimens and based on the number of individuals in TB compartment 6 (active TB). The costs for treatment for PLWH include inpatient and outpatient treatments for causes other than TB. They are differentiated by ART status and CD4 count, and are based on the number of PLWH in HIV compartment 2

(HIV+, not on ART, CD4>200), HIV compartment 3 (HIV+, not on ART, CD4 $\leq$ 200), and HIV compartment 4 (HIV+, on ART) and not in TB compartment 6 (without active TB). We calculate the cost of HIV testing based on the number of individuals initiating ART and the estimated cost of finding one person to initiate ART. The cost model equations for each care program is described in Section 4.5.

##### 1.4 Incremental Health Outcomes and Costs

We quantify discounted and undiscounted health outcomes, including TB incident cases, TB deaths, and DALYs, and costs for each program over the 10-year intervention period. We use these metrics to generate incremental cost-effectiveness ratios (ICERs) to assess the undiscounted and discounted per-dollar cost per incident TB case averted, TB death averted, and DALYs averted of community-based care programs compared to facility-based care programs, including:

- Program 2 (community-based ART care with standard facility-based TPT care) to Program 1 (standard facility-based ART and TPT care) to quantify incremental health gains and additional costs of a community-based ART intervention,
- Program 3 (community-based ART with TPT care) compared to Program 1 (standard facility-based ART and TPT care) to quantify incremental health gains and additional costs of a community-based ART and TPT intervention, and
- Program 3 (community-based ART with TPT care) compared to Program 2 (community-based ART care with standard facility-based TPT care) to quantify incremental health gains and additional costs of a nested community-based TPT intervention.

The equations used to generate ICERs are described in Section 4.6. We use discounted ICERs to evaluate the cost-effectiveness of programs in the manuscript.

#### 2 Dynamic Transmission Model Equations

Time-varying parameters that are calculated based on current population states at each time step $\tau$, including TB force of infection calculations, ART initiations, and population entries and exits, are governed by the equations described in Section 2.1. Rates of flow between compartments are governed by the system of ordinary differential equations defined in Section 2.2.

##### 2.1 Time-Varying Parameters Impacted by Population States

A description of the parameters used in the calculations of TB force of infection calculations, ART initiation rates, and entries to and exits from the population is given in Table 2. These parameters are calculated based on the total population in TB compartment $t$, DR compartment $r$, HIV compartment $h$, and gender compartment $g$ at time $\tau$ denoted $N_{t,r,h,g}(\tau)$.

###### 2.1.1 TB Force of Infection

The TB force of infection for DR compartment $r$ at time $\tau$ is denoted $\lambda_r(\tau)$, and is calculated at each time step using Equations (A) through (C). The number of individuals infected with active DS-TB (in HIV compartment 6 and DR compartment 1), HIV compartment $h$, and gender $g$ at time $\tau$ is indicated by $N_{6,1,h,g}(\tau)$. Relative transmissibility varies by HIV compartment $h$, and is represented by the parameter $\phi_h$ [21, 30]. The estimated fraction of new TB infections that are

| Notation | Description |
|---|---|
| **Parameters that impact TB force of infection ($\lambda_r(\tau)$)** | |
| $\beta_g$ | Number of effective contacts for TB transmission per year for gender $g$ |
| $\phi_h$ | Relative transmissibility of TB in populations in HIV compartment $h$, such that $\phi_1 = 1$ and $\phi_h < 1$ for $h = 2, 3$, or 4 |
| $\varepsilon$ | Fraction of new TB infections that are MDR-TB |
| **Parameters that impact ART initiation ($\eta_{2,4,g}(\tau)$ and $\eta_{3,4,g}(\tau)$)** | |
| $\sigma_g(\tau)$ | ART coverage for gender $g$ at time $\tau$. |
| $\varrho_g(\tau)$ | Proportion of the population in HIV compartment 2 (HIV+, not on ART, CD4 > 200) eligible to initiate ART by gender $g$ at time $\tau$. |
| **Parameters that impact entries ($B(\tau)$) and exits ($\alpha^{out}_{t,h,g}(\tau)$) from the population** | |
| $\mu_{t,h,g}(\tau)$ | Mortality rates from populations in TB compartment $t$, HIV compartment $h$, and gender compartment $g$ at time $\tau$, per year |
| $\alpha^{\text{ageout}}$ | Rate of exit from the population due to aging, $\alpha^{\text{ageout}} = 1/(60 - 15)$ |

Table 2: Description and notation for parameters used in calculations impacted by population states.

projected to be MDR are represented by the parameter $\varepsilon$. These parameters are used to calculate the TB force of infection for each DR compartment $r$ at each time step $\tau$, as follows

$$\lambda_{1,g}(\tau) = \left(\frac{\beta_g}{100,000}\right) \sum_{h \in HIV} \phi_h N_{6,1,h,g}(\tau) \qquad \forall g \in G \qquad \text{(A)}$$

$$\lambda_{2,g}(\tau) = \frac{\varepsilon \lambda_{1,g}(\tau)}{(1-\varepsilon)} \qquad \forall g \in G \qquad \text{(B)}$$

$$\lambda_r(\tau) = \sum_{g \in G} \lambda_{r,g}(\tau) \qquad \forall r \in DR. \qquad \text{(C)}$$

Equation (A) calculates the TB force of infection for DS-TB ($r = 1$) for each gender $g$. Equation (B) uses the estimated fraction of new TB infections that are projected to be MDR to calculate the TB force of infection for MDR-TB strains ($r = 2$) by gender $g$. Finally, Equation (C) sums over gender $g$ to get the total TB force of infection for each DR compartment $r$.

##### 2.1.2 ART Initiation Rates

ART initiation rates ($\eta_{2,4,g}(\tau)$ and $\eta_{3,4,g}(\tau)$) are estimated based on: ART coverage by gender $g$ at time $\tau$, denoted $\sigma_g(\tau)$; the proportion of those who are in HIV compartment 2 (HIV+, not on ART, CD4 > 200) and are eligible to initiate ART by gender $g$ at time $\tau$, denoted $\varrho_g(\tau)$; and the current proportion of those who are HIV positive in each HIV compartment $h \in HIV\{2, 3, 4\}$, by gender $g$ at time $\tau$, denoted $V_{h,g}(\tau)$. ART coverage, $\sigma_g(\tau)$, is used to calculate ART initiation rates so that the proportion of PLWH on ART by gender $g$ corresponds to ART coverage assumptions. There are four distinct time periods that impact the input parameter values for ART coverage, $\sigma_g(\tau)$, and the proportion of those in HIV compartment 2 (HIV+, not on ART, CD4 > 200) that are eligible to initiate ART, $\varrho_g(\tau)$ [37],

1. Before 2004: ART was not available in this setting.
2. Between 2004-2010: Individuals living with HIV with CD4 $\leq$ 200 were eligible for ART.
3. Between 2011-2015: CD4 eligibility levels were increased to 350 for all individuals who were HIV positive.
4. After 2016: All individuals who are HIV positive are eligible for ART.

The input values for ART coverage at time $\tau$, by gender $g$, $\sigma_g(\tau)$ and the proportion of those in HIV compartment 2 (HIV+, not on ART, CD4 $>$ 200) that are eligible to initiate ART by gender, $\varrho_g(\tau)$, reflect these time periods. The values for ART coverage over time, by gender, $\sigma_g(\tau)$, are described in Section 5.2.3 and Section 5.3.1. The proportion of those in HIV compartment 2 (HIV+, not on ART, CD4 $>$ 200) that are eligible to initiate ART by gender, $\varrho_g(\tau)$, is described in Section 5.2.4. We assume all those eligible to initiate ART, initiate at the same rate.

Before 2004, all ART initiation rates are set to zero, i.e. $\eta_{2,4,g}(\tau) = \eta_{3,4,g}(\tau) = 0$, to represent that ART is not available before 2004. After 2004, ART initiation rates are calculated with input parameters and Equation (D), Equation (E), Equation (F), and Equation (G). Equation (D) calculates the proportion of HIV positive population by gender $g$ in the model in each HIV compartment $h$ at time $\tau$, $V_{h,g}(\tau)$, given as

$$V_{h,g}(\tau) = \frac{\sum_{t \in TB} \sum_{r \in DR} N_{t,r,h,g}(\tau)}{\sum_{t \in TB} \sum_{r \in DR} \sum_{\substack{i \in HIV \\ \{2,3,4\}}} N_{t,r,i,g}(\tau)} \qquad \forall h \in HIV\{2,3,4\}, g \in G. \tag{D}$$

Equation (E) calculates a general ART initiation rate for all PLWH, not on ART (in HIV compartment 2 and HIV compartment 3) eligible to initiate ART by gender, denoted $\eta_g^{all}(\tau)$ (assuming that all those eligible to initiate ART, initiate at the same rate). The numerator represents the difference in ART coverage for gender $g$ at time $\tau$ with $\sigma_g(\tau)$ and model projected ART coverage for gender $g$ at time $\tau$, $V_{4,g}(\tau)$. The denominator represents the proportion of PLWH, not on ART (in HIV compartment 2 and HIV compartment 3) eligible to initiate ART at time $\tau$ by gender $g$. The general ART initiation rate is calculated as

$$\eta_g^{all}(\tau) = \frac{\sigma_g(\tau) - V_{4,g}(\tau)}{\varrho_g(\tau) V_{2,g}(\tau) + V_{3,g}(\tau)} \qquad \forall g \in G. \tag{E}$$

Since all those in HIV compartment 2 (HIV+, not on ART, CD4 $>$ 200) are eligible to initiate ART for all times associated with the years after 2004, Equation (F) sets ART initiation rates for those in HIV compartment 2, $\eta_{2,4,g}(\tau)$, to the general ART initiation rate for all PLWH, not on ART $\eta_g^{all}(\tau)$ as

$$\eta_{2,4,g}(\tau) = \eta_g^{all}(\tau) \qquad \forall g \in G. \tag{F}$$

Finally, Equation (G) provides ART initiation rates from HIV compartment 3 (HIV+, not on ART, CD4 $\leq$ 200) to HIV compartment 4 (HIV+, on ART), by gender at time $\tau$ as

$$\eta_{3,4,g}(\tau) = \varrho_g(\tau) \eta_g^{all}(\tau) \qquad \forall g \in G. \tag{G}$$

##### 2.1.3 Population Entering and Exiting the Model

This section describes how we calculate the rate of population exiting the model based on aging out rates and mortality rates at time $\tau$, $\alpha^{out}_{t,h,g}(\tau)$ for TB compartment $t$, DR compartment $r$, HIV compartment $h$ and gender compartment $g$ in Equation (H), and the total population aging into the model at time $\tau$, $B(\tau)$ in Equation (I).

Exits from the model can occur due to mortality at a rate of $\mu_{t,h,g}(\tau)$ for TB compartment $t$, HIV compartment $h$ and gender compartment $g$ at time $\tau$ or aging out at a rate of $\alpha^{\text{ageout}}$. We only consider populations with ages between 15 and 59, that is, they enter on their 15th birthday and exit on their 60th birthday so we assume aging out of the population occurs at a rate of $1/(60-15)$ per year which is the inverse of the duration of time between population entry at age 15 and exit at age 60. So, we set $\alpha^{\text{ageout}} = 1/(60-15)$. Note that $\mu_{t,h,g}(\tau)\alpha^{\text{ageout}}$ is subtracted to ensure an individual can only age out or die, but cannot both age out and die. For the purpose of this model, we assume a constant population size of 100,000. In order to keep the population size constant, the entries to the population are set to the total population leaving the model from aging out or dying. The rate of population exiting the model based on aging out rates and mortality rates are calculated as

$$\alpha^{out}_{t,h,g}(\tau) = \mu_{t,h,g}(\tau) + \alpha^{\text{ageout}}(1 - \mu_{t,h,g}(\tau)) \qquad \forall t \in TB, h \in HIV, g \in G. \tag{H}$$

The total population aging into the model is calculated as

$$B(\tau) = \sum_{t\in TB}\sum_{r\in DR}\sum_{h\in HIV}\sum_{g\in G}\alpha^{out}_{t,h,g}(\tau)N_{t,r,h,g}(\tau). \tag{I}$$

New entries are assigned to TB uninfected, latent TB, and active TB compartments, $t \in TB\{1,3,6\}$, HIV- and HIV+, not on ART, CD4 > 200, $h \in HIV\{1,2\}$, as well as all DR $r$ compartments and gender $g$ compartments at time $\tau$ with the input parameter $\alpha^{in}_{t,r,h,g}(\tau)$.

#### 2.2 Model Transitions

This section describes the differential equations used to represent transitions between the eight TB compartments, two DR compartments, four HIV compartments, and two gender compartments.

*TB compartment 1: Uninfected, not on TPT**

$$\begin{aligned}\frac{dN_{1,1,h,g}(\tau)}{dt} = \;& \alpha^{in}_{1,1,h,g}(\tau)B(\tau) \\ & - \alpha^{out}_{1,h,g}(\tau)N_{1,1,h,g}(\tau) \\ & - \kappa_{h,g}(\tau)N_{1,1,h,g}(\tau) \\ & + \omega N_{2,1,h,g}(\tau) \\ & - \sum_{r\in DR}\lambda_r(\tau)N_{1,1,h,g}(\tau) \\ & + \sum_{i\in HIV, i\neq h}\eta_{i,h,g}(\tau)N_{1,1,i,g}(\tau) \\ & - \sum_{i\in HIV, i\neq h}\eta_{h,i,g}(\tau)N_{1,1,h,g}(\tau)\end{aligned} \tag{1}$$

$$\forall h \in HIV, g \in G$$

**Note: we do not distinguish TB uninfected, not on TPT, compartments by drug resistance, and as such, all initial uninfected populations are assigned to DR compartment 1, and all exits and entries into TB uninfected compartments are from DR compartment 1.*

*Equation* (1) *Description*
The first line in Equation (1) calculates the entries into TB compartment 1 as a proportion of the population enters at age 15. The second line calculates the population leaving the compartment from aging out or dying. The third and fourth lines calculate the total population leaving and entering compartment 1 from initiation (only those who are HIV+ and on ART) and completion of TPT, respectively. The fifth line calculates the population leaving after being infected with TB. The last two lines calculate entries and exits between HIV compartments.

*TB compartment 2: Uninfected, on TPT**

$$
\begin{aligned}
\frac{dN_{2,1,h,g}(\tau)}{dt} = \quad & -\alpha^{out}_{2,h,g}(\tau)N_{2,1,h,g}(\tau) \\
& +\kappa_{h,g}(\tau)N_{1,1,h,g}(\tau) \\
& -\omega N_{2,1,h,g}(\tau) \\
& -\sum_{r\in DR}\iota_r\lambda_r(\tau)N_{2,1,h,g}(\tau) \\
& +\sum_{i\in HIV, i\neq h}\eta_{i,h,g}(\tau)N_{2,1,i,g}(\tau) \\
& -\sum_{i\in HIV, i\neq h}\eta_{h,i,g}(\tau)N_{2,1,h,g}(\tau) \qquad (2) \\
& \forall h\in HIV, g\in G
\end{aligned}
$$

**Note: We do not distinguish TB uninfected, not on TPT, compartments by drug resistance, and as such, all initial uninfected populations are assigned to DR compartment 1, and all exits and entries into TB uninfected compartments are from DR compartment 1.*

*Equation* (2) *Description*
The first line in Equation (2) calculates the population leaving the compartment from aging out or dying (note: we do not let individuals age into TPT compartments). The second and third lines calculate the total population entering and leaving compartment 2 from initiation and completion of TPT. The fourth line calculates the population leaving after being infected with TB, diminished for those infected with DS TB to represent the partially protective effects of becoming infected with TB while on TPT. The last two lines calculate entries and exits between HIV compartments.

*TB compartment 3: LTBI, infected recently*

$$
\begin{aligned}
\frac{dN_{3,r,h,g}(\tau)}{dt} = \quad & \alpha^{in}_{3,r,h,g}(\tau)B(\tau) \\
& -\alpha^{out}_{3,h,g}(\tau)N_{3,r,h,g}(\tau) \\
& +\lambda_r(\tau)N_{1,1,h,g}(\tau) \\
& +\iota_r\lambda_r(\tau)N_{2,1,h,g}(\tau)
\end{aligned}
$$

$$
\begin{aligned}
&+ \zeta\lambda_r(\tau) \sum_{r \in DR} N_{4,r,h,g}(\tau) \\
&+ \xi\lambda_r(\tau) \sum_{r \in DR} N_{7,r,h,g}(\tau) \\
&+ \zeta\lambda_r(\tau) \sum_{r \in DR} N_{8,r,h,g}(\tau) \\
&- \pi_{3,4} N_{3,r,h,g}(\tau) \\
&- \gamma_r \kappa_{h,g}(\tau) N_{3,r,h,g}(\tau) \\
&- \theta_h \pi_{3,6} N_{3,r,h,g}(\tau) \\
&+ \sum_{i \in HIV, i \neq h} \eta_{i,h,g}(\tau) N_{3,r,i,g}(\tau) \\
&- \sum_{i \in HIV, i \neq h} \eta_{h,i,g}(\tau) N_{3,r,h,g}(\tau) \qquad (3)
\end{aligned}
$$

$$\forall r \in DR, h \in HIV, g \in G$$

*Equation* (3) *Description*

Line one in Equation (3) calculates entries into TB compartment 3 as a proportion of the population enters at age 15. Line two calculates the population leaving the compartment from aging out or dying. Lines three through seven calculate entries from infections (and re-infections) from TB compartments 1 (uninfected, not on TPT), 2 (uninfected, on TPT), 4 (LTBI, remote), 8 (LTBI, on TPT), and 7 (recovered/treated). For the purpose of this model, we assume in regards to reinfection, that the most recent TB strain type determines the DR compartment. As such, we sum the total populations from compartments 4, 7, and 8 over drug-resistant compartments to allow populations to transition between DR compartments (DS and MDR). For individuals in TB compartments 4 and 8, re-infection is diminished by the partially protective effect of previous infections on reinfection. For individuals in TB compartment 8, infection rates are increased to account for the increased risk of reinfection after active TB. Lines eight through ten calculate exits due to movements from recent to remote, going onto TPT, and progression to active TB. We only allow individuals with DS LTBI and HIV+ and on ART to move into TB compartment 5, we assume those with MDR LTBI on TPT continue to progress according to the annual risk of progression for recent LTBI. The last two lines calculate transitions between HIV compartments for entries and exits.

*TB compartment 4: LTBI, infected remotely*

$$
\begin{aligned}
\frac{dN_{4,r,h,g}(\tau)}{dt} = \;& \alpha^{in}_{4,r,h,g}(\tau) B(\tau) \\
&- \alpha^{out}_{4,h,g}(\tau) N_{4,r,h,g}(\tau) \\
&+ \pi_{3,4} N_{3,r,h,g}(\tau) \\
&- \zeta\lambda_r(\tau) N_{4,r,h,g}(\tau) \\
&- \gamma_r \kappa_{h,g}(\tau) N_{4,r,h,g}(\tau) \\
&- \theta_h \pi_{4,6} N_{4,r,h,g}(\tau) \\
&+ \sum_{i \in HIV, i \neq h} \eta_{i,h,g}(\tau) N_{4,r,i,g}(\tau) \\
&- \sum_{i \in HIV, i \neq h} \eta_{h,i,g}(\tau) N_{4,r,h,g}(\tau) \qquad (4)
\end{aligned}
$$

$$\forall r \in DR, h \in HIV, g \in G$$

*Equation* (4) *Description*
Line one in Equation (4) calculates entries into TB compartment 4 as a proportion of the population enters at age 15. Line two calculates the population leaving the compartment from aging out or dying. Line three calculates entries from LTBI, recent to remote, two years after the initial LTBI. Lines three to six calculate exits due to re-infection, which is diminished for the partially-protective effect of previous LTBI infections against acquiring a new TB infection, going onto TPT, and progression to active TB. We only allow individuals with DS LTBI and HIV+ and on ART to move into TB compartment 5. We assume those with MDR LTBI on TPT continue to progress according to the annual risk of progression for remote LTBI. The last two lines calculate transitions between HIV compartments.

*TB compartment 5: LTBI, on TPT*

$$\begin{aligned}\frac{dN_{5,r,h,g}(\tau)}{dt} = \quad & -\alpha^{out}_{5,h,g}(\tau)N_{5,r,h,g}(\tau) \\ & + \gamma_r \kappa_{h,g}(\tau)(N_{3,r,h,g}(\tau) + N_{4,r,h,g}(\tau)) \\ & - \pi_{5,6}N_{5,r,h,g}(\tau) \\ & - \omega N_{5,r,h,g}(\tau) \\ & + \sum_{i \in HIV, i \neq h} \eta_{i,h,g}(\tau)N_{5,r,i,g}(\tau) \\ & - \sum_{i \in HIV, i \neq h} \eta_{h,i,g}(\tau)N_{5,r,h,g}(\tau) \end{aligned} \tag{5}$$

$$\forall r \in DR, h \in HIV, g \in G$$

*Equation* (5) *Description*
Line one in Equation (5) calculates the population leaving the compartment from aging out or dying (note: we do not let individuals age into TPT compartments). Line two calculates the rate at which populations move onto TPT from the LTBI, infected recently, and LTBI, infected remotely compartments, respectively. Lines three and four calculate exits due to active TB progression and completion of TPT. The last two lines calculate transitions between HIV compartments for entries and exits.

*TB compartment 6: Active TB*

$$\begin{aligned}\frac{dN_{6,r,h,g}(\tau)}{dt} = \quad & \alpha^{in}_{6,r,h,g}(\tau)B(\tau) \\ & - \alpha^{out}_{6,h,g}N_{6,r,h,g}(\tau) \\ & + \theta_h \pi_{3,6}N_{3,r,h,g}(\tau) \\ & + \theta_h \pi_{4,6}N_{4,r,h,g}(\tau) \\ & + \pi_{5,6}N_{5,r,h,g}(\tau) \\ & + \pi_{8,6}N_{8,r,h,g}(\tau) \\ & + \pi_{7,6}N_{7,r,h,g}(\tau) \end{aligned}$$

$$
\begin{aligned}
& - \upsilon_h \pi_{6,7} N_{6,r,h,g}(\tau) \\
& + \sum_{i \in HIV, i \neq h} \eta_{i,h,g}(\tau) N_{6,r,i,g}(\tau) \\
& - \sum_{i \in HIV, i \neq h} \eta_{h,i,g}(\tau) N_{6,r,h,g}(\tau) \qquad (6)
\end{aligned}
$$

$$\forall r \in DR, h \in HIV, g \in G$$

*Equation* (6) *Description*
Line one in Equation (6) calculates entries into TB compartment 6 as a proportion of the population enters at age 15. Line two calculates the population leaving the compartment from aging out or dying. Lines three through six calculate the rate at which populations with LTBI who are recently and remotely infected (relative to HIV state), on TPT, and after TPT (for those who are HIV+ and on ART) progress to active TB. Line seven calculates entries from relapse. Line eight calculates exits from recovery and accounts for the increased duration of active TB due to delays in treatment. The last two lines calculate transitions between HIV compartments for entries and exits.

*TB compartment 7: Recovered/Treated*

$$
\begin{aligned}
\frac{dN_{7,r,h,g}(\tau)}{dt} = \quad & - \alpha^{out}_{7,h,g} N_{7,r,h,g}(\tau) \\
& + \upsilon_h \pi_{6,7} N_{6,r,h,g}(\tau) \\
& - \xi \lambda_r \sum_{r \in DR} N_{7,r,h,g}(\tau) \\
& - \pi_{7,6} N_{7,r,h,g}(\tau) \\
& + \sum_{i \in HIV, i \neq h} \eta_{i,h,g}(\tau) N_{7,r,i,g}(\tau) \\
& - \sum_{i \in HIV, i \neq h} \eta_{h,i,g}(\tau) N_{7,r,h,g}(\tau) \qquad (7)
\end{aligned}
$$

$$\forall r \in DR, h \in HIV, g \in G$$

*Equation* (7) *Description*
Line one in Equation (7) calculates the population leaving the compartment from aging out or dying (note: we do not let individuals age into the TB recovered/treated compartment). Line two calculates entries from recovery/treatment and accounts for the increased duration of active TB due to delays in treatment. Line three calculates exits from reinfection, which accounts for the increased risk of acquiring a new TB infection after active TB. Line four calculates exits from relapse. The last two lines calculate transitions between HIV compartments.

*TB compartment 8: LTBI after TPT*

$$
\begin{aligned}
\frac{dN_{8,r,h,g}(\tau)}{dt} = \quad & - \alpha^{out}_{8,h,g} N_{8,r,h,g}(\tau) \\
& + \omega N_{5,r,h,g}(\tau) \\
& - \pi_{8,6} N_{8,r,h,g}(\tau)
\end{aligned}
$$

$$
\begin{aligned}
&- \zeta\lambda(\tau) \sum_{r \in DR} N_{8,r,h,g}(\tau) \\
&+ \sum_{i \in HIV, i \neq h} \eta_{i,h,g}(\tau) N_{8,r,i,g}(\tau) \\
&- \sum_{i \in HIV, i \neq h} \eta_{h,i,g}(\tau) N_{8,r,h,g}(\tau) \qquad (8)
\end{aligned}
$$

$$\forall r \in DR, h \in HIV, g \in G$$

*Equation* (8) *Description*
Line one in Equation (8) calculates the population leaving the compartment from aging out or dying (note: we do not let individuals age into TPT compartments). Line two calculates the total population entering TB compartment 8 after completing TPT with LTBI. Lines three and four calculate exits due to progression to active TB and reinfection, which is diminished for the partially-protective effect of LTBI from TB against acquiring a new TB infection. The last two lines calculate transitions between HIV compartments for entries and exits.

### 3 Model Execution

The dynamic transmission model was programmed in R version 3.5.2 and the system of differential equations was solved with the deSolve package [42]. We run our code on Hyak, the University of Washington's supercomputing system, to allow for computations at scale [22]. The code and parameters used to run the model are available at `https://github.com/cgreene3/epi_model_HIV_TB_KZN_SA/tree/master/model_execution`. We start the dynamic transmission model at the beginning of 1940 at time $\tau = 1940.0$ to allow for the slow propagation of TB. We initiate population states at the start of 1940 into TB compartments 1 (Uninfected, not on TPT), 3 (LTBI, recent), 4 (LTBI, remote), and 6 (active TB), DR compartment 1 (DS-TB), HIV compartment 1 (HIV negative), and both gender compartments 1 and 2. We introduce HIV through HIV incidence rates at the beginning of 1980 at time $\tau = 1980.0$.

We solve the system of differential equations using a time step $\tau$ of one month or 1/12 (0.083) of a year. Each time step $\tau$ is associated with a year, *year y*. For example time steps $\tau \in \{1990.0, 1990.083, \ldots, 1990.833, 1990.917\}$ are associated with the year 1990 or *year* $y = 1990$. Time-varying parameters are notated as a function of $\tau$ in Table 1.

To calibrate parameters used in the dynamic transmission model, we generate a set of parameter sets from a range of values as described in Section 5 that correspond to findings from the DO ART trial and scientific literature for each parameter included in the calibration. We evaluate the model outputs for each parameter set against calibration target ranges as described in Section 5.4 to determine whether to accept or reject a parameter set. The source code to generate the set of parameter sets can be found at `https://github.com/cgreene3/epi_model_HIV_TB_KZN_SA/blob/master/param_files/calculated_param_gen/9_sample_gen.R`. The source code for the model for the warm-up and calibration period is available at `https://github.com/cgreene3/epi_model_HIV_TB_KZN_SA/blob/master/model_execution/calibration_runs/warmup_calibration_for_loop_Rscript.R`. The script used to generate the set of accepted parameter sets can be found at `https://github.com/cgreene3/epi_model_HIV_TB_KZN_SA/blob/master/model_execution/calibration_runs/calibration_analysis_Rscript.R`. The results from the calibration can be found at `https://github.com/cgreene3/epi_model_HIV_TB_KZN_SA/blob/master/results/calibration_analysis/`.

Then, we continue to execute the model for the three care programs and each of the accepted parameter sets over the intervention period, from the beginning of 2018 until the end of 2027.

The population states at the end of 2017 for each accepted parameter set are used to initiate the population states for each of the three care programs at the beginning of 2018. We modify parameters that depend on the care program starting at the beginning of 2018 and hold them constant over the intervention period. The source code for the model intervention period is available at `https://github.com/cgreene3/epi_model_HIV_TB_KZN_SA/blob/master/model_execution/program_runs/program_runs_for_loop_Rscript.R`.

We calculate program health outcome metrics for each parameter set and care program based on the equations in Section 4. The code used to generate program health outcomes and graphs can be found at `https://github.com/cgreene3/epi_model_HIV_TB_KZN_SA/blob/master/model_execution/program_runs/program_analysis_Rscript.R`.

### 4 Program Health Outcomes and Costs

Program health outcomes and costs are summarized in Table 3. The metrics related to TB incidence, TB mortality, TB and HIV prevalence, and DALYs, are based on population states and transition rates from the dynamic transmission model. The equations used to generate these metrics are described in Sections 4.1 through 4.4. The cost model generates costs for each program, as described in Section 4.5. We also calculate discounted and undiscounted program health outcomes and costs over the 10-year intervention period from the start of 2018 to the end of 2027, as described in Section 4.6. Discounted and undiscounted incremental cost-effectiveness ratios per TB incidence, TB deaths, and DALYs averted between the care programs are also described in Section 4.6.

| **Notation** | **Description** |
|---|---|
| | **TB Incidence** |
| $TBinc\_per(y,p)$ | TB incidence rate in *year y*, per 100,000 individuals for program $p$ |
| $TBinc\_per_g^{HIV+}(y,p)$ | TB incidence rate for the HIV positive population in *year y* by gender $g$, per 100,000 males or females for program $p$ |
| $TBinc\_per_g^{HIV-}(y,p)$ | TB incidence rate for the HIV negative population in *year y* by gender $g$, per 100,000 males or females for program $p$ |
| | **TB Mortality** |
| $TBmort\_per(y,p)$ | TB mortality rate in *year y*, per 100,000 individuals for program $p$ |
| $TBmort\_per_g^{HIV+}(y,p)$ | TB mortality rate for the HIV positive population in *year y* by gender $g$, per 100,000 males or females for program $p$ |
| $TBmort\_per_g^{HIV-}(y,p)$ | TB mortality rate for the HIV negative population in *year y* by gender $g$, per 100,000 males or females for program $p$ |
| | **TB and HIV prevalence** |
| $TBprev\_per(y,p)$ | TB prevalence rate in *year y*, per 100,000 individuals for program $p$ |
| $HIVprev\_per_g(y,p)$ | HIV prevalence rate in *year y* by gender $g$, per 100,000 males or females for program $p$ |
| | **Disability-Adjusted Life Years (DALYs)** |
| $YLL\_per(y,p)$ | YLL in *year y*, per 100,000 individuals for program $p$ |
| $YLD\_per(y,p)$ | YLD in *year y*, per 100,000 individuals for program $p$ |
| $DALY\_per(y,p)$ | DALYs in *year y*, per 100,000 individuals for program $p$ |
| | **Costs** |
| $Cost\_per(y,p)$ | Costs in *year y*, per 100,000 individuals for program $p$ |
| | Continued on next page |

**Table 3 – continued from previous page**

| Notation | Description |
|---|---|
| **Program Health Outcomes and Costs (Undiscounted)** | |
| $UTBinc\_total(p)$ | Total undiscounted incident TB cases over the 10-year intervention period from 2018 to 2027 for program $p$ |
| $UTBmort\_total(p)$ | Total undiscounted TB deaths over the 10-year intervention period from 2018 to 2027 for program $p$ |
| $UDALY\_total(p)$ | Total undiscounted DALYs over the 10-year intervention period from 2018 to 2027 for program $p$ |
| $UCost\_total(p)$ | Total undiscounted costs over the 10-year intervention period from 2018 to 2027 for program $p$ |
| **Program Health Outcomes and Costs (Discounted)** | |
| $DTBinc\_total(p)$ | Total discounted incident TB cases over the 10-year intervention period from 2018 to 2027 for program $p$ |
| $DTBmort\_total(p)$ | Total discounted TB deaths over the 10-year intervention period from 2018 to 2027 for program $p$ |
| $DDALY\_total(p)$ | Total discounted DALYs over the 10-year intervention period from 2018 to 2027 for program $p$ |
| $DCost\_total(p)$ | Total discounted costs over the 10-year intervention period from 2018 to 2027 for program $p$ |
| **Incremental Cost-Effectiveness Ratios (Undiscounted)** | |
| $UICER^{TBinc}(\tilde{p}, p)$ | Undiscounted incremental cost per TB incidence case averted from 2018 to 2027 between the intervention program $\tilde{p}$ and program $p$ |
| $UICER^{TBmort}(\tilde{p}, p)$ | Undiscounted incremental cost per TB death averted from 2018 to 2027 between the intervention program $\tilde{p}$ and program $p$ |
| $UICER^{DALY}(\tilde{p}, p)$ | Undiscounted incremental cost per DALY from 2018 to 2027 between the intervention program $\tilde{p}$ and program $p$ |
| **Incremental Cost-Effectiveness Ratios (Discounted)** | |
| $DICER^{TBinc}(\tilde{p}, p)$ | Discounted incremental cost per TB incidence case averted over the intervention period between the intervention program $\tilde{p}$ and program $p$ |
| $DICER^{TBmort}(\tilde{p}, p)$ | Discounted incremental cost per TB death averted over the intervention period between the intervention program $\tilde{p}$ and program $p$ |
| $DICER^{DALY}(\tilde{p}, p)$ | Discounted incremental cost per DALY averted over the intervention period between the intervention program $\tilde{p}$ and program $p$ |

Table 3: Description and notation for program health outcomes and costs.

Metrics that are representative of rates (per 100,000 individuals, per 100,000 males, or per 100,000 females) contain *per* in the notation. Estimates that are cumulative over the 10-year intervention period contain *total* in the notation. Metrics are notated as a function of $p$ for Programs $p = 1, p = 2$, and $p = 3$. Annual rates, notated as a function of $y$, are generated by summing model projections over time steps $\tau$ associated with *year* $y$, where $y$ takes on values from 2018 to 2027. Since we consider monthly time steps, there are 12 values of $\tau$ in each year. For example, the annual rate for *year* $y = 2018$ sums over time steps $\tau \in \{2018.0, 2018.083, \ldots, 2018.833, 2018.917\}$. We generate estimates that are cumulative over the 10-year intervention period by adding all yearly rates associated with the years in the intervention period, *year* $y \in [2018, 2027]$.

Rates that are representative per 100,000 males or per 100,000 females contain a subscript of $g$ ($g = 1$ for males and $g = 2$ for females). TB incidence rates and TB mortality with a superscript of $HIV+$ aggregate those in HIV positive compartments (HIV compartments 2, 3, and 4), and a superscript of $HIV-$ represents the HIV negative population (HIV compartment 1). Discounted and undiscounted incremental cost-effectiveness ratios (ICERs) compare a community-based intervention program $\tilde{p}$ to program $p$.

##### 4.1 TB Incidence

TB incidence rates per 100,000 individuals are calculated by summing those progressing to active TB (TB compartment 6) from TB compartments 3 (LTBI, recent), 4 (LTBI, remote), 5 (LTBI, on TPT), and 8 (LTBI, after TPT). Note: only those in HIV compartment 4 (HIV+, and on ART) can initiate TPT and enter TB compartments 5 and 8. TB progression rates from TB compartments 5 and 8 are defined for PLWH on ART. We sum over all time steps $\tau$ in *year y* to calculate TB incidence rates for *year y* for program $p$ as,

$$\begin{aligned} TBinc\_per(y,p) = &\sum_{\tau \in year\ y} \sum_{\substack{t \in TB \\ \{3,4\}}} \sum_{r \in DR} \sum_{h \in HIV} \sum_{g \in G} \theta_h \pi_{t,6} N_{t,r,h,g}(\tau) \\ &+ \sum_{\tau \in year\ y} \sum_{\substack{t \in TB \\ \{5,8\}}} \sum_{r \in DR} \sum_{g \in G} \pi_{t,6} N_{t,r,4,g}(\tau). \end{aligned} \tag{9}$$

We also generate TB incidence rates by year and by gender (per 100,000 males or females) for those who are HIV positive and HIV negative in Equation (11) and Equation (12), respectively. In order to scale rates by gender, we use population estimates for each gender compartment $g$ at time $\tau$ for program $p$, denoted $POP_g(\tau,p)$. Population estimates for gender compartment $g$ at time $\tau$ for program $p$ is calculated as,

$$POP_g(\tau,p) = \sum_{t \in TB} \sum_{r \in DR} \sum_{h \in HIV} N_{t,r,h,g}(\tau) \qquad \forall g \in G. \tag{10}$$

Equation (11) calculates the number of TB incident cases for the HIV positive population by gender $g$ at time $\tau$, then scales TB incidence rates for those who are HIV positive per 100,000 males $g = 1$ and 100,000 females $g = 2$ by dividing by population estimates for each gender $g$, summing over all time steps $\tau$ in *year y* and multiplying by 100,000,

$$\begin{aligned} TBinc\_per_g^{HIV+}(y,p) = &\ 100,000 \sum_{\tau \in year\ y} \left( \frac{\sum_{\substack{t \in TB \\ \{3,4\}}} \sum_{r \in DR} \sum_{\substack{h \in HIV \\ \{2,3,4\}}} \theta_h \pi_{t,6} N_{t,r,h,g}(\tau)}{POP_g(\tau,p)} \right) \\ &+ 100,000 \sum_{\tau \in year\ y} \left( \frac{\sum_{\substack{t \in TB \\ \{5,8\}}} \sum_{r \in DR} \pi_{t,6} N_{t,r,4,g}(\tau)}{POP_g(\tau,p)} \right) \qquad \forall g \in G. \end{aligned} \tag{11}$$

Similarly, Equation (12) calculates the number of TB incident cases for the HIV negative population by gender $g$ at time $\tau$ in program $p$, and divides by population estimates for each gender $g$, then summing over all time steps $\tau$ in *year y* and multiplying by 100,000,

$$TBinc\_per_g^{HIV-}(y,p) = 100,000 \sum_{\tau \in year\ y} \left( \frac{\sum\limits_{\substack{t \in TB \\ \{3,4\}}} \sum\limits_{r \in DR} \pi_{t,6}\ N_{t,r,1,g}(\tau)}{POP_g(\tau,p)} \right) \qquad \forall g \in G. \tag{12}$$

The aggregated TB incidence rate in *year* $y$, per 100,000 males ($g = 1$) and females ($g = 2$) for program $p$, is equal to the sum of $TBinc\_per_g^{HIV+}(y,p)$ plus $TBinc\_per_g^{HIV-}(y,p)$ by gender $g$.

#### 4.2 TB Mortality

TB mortality per 100,000 individuals projected by the model for program $p$ is calculated by summing those departing the active TB compartment (TB compartment 6) due to death for all time steps $\tau$ in *year* $y$ and is calculated as,

$$TBmort\_per(y,p) = \sum_{\tau \in year\ y} \sum_{r \in DR} \sum_{h \in HIV} \sum_{g \in G} \mu_{6,h,g}(\tau)\ N_{6,r,h,g}(\tau). \tag{13}$$

We also generate TB mortality in program $p$ by gender (per 100,000 males or females) for those who are HIV positive and HIV negative in Equation (14) and Equation (15), respectively. We use $POP_g(\tau,p)$ as described in Equation (10) to scale TB mortality by gender. Equation (14) calculates the number of HIV positive individuals who died at time $\tau$, divided by the number of individuals who are HIV positive by population estimates for each gender $g$, and summed over all time steps $\tau$ in *year* $y$ and multiplied by 100,000,

$$TBmort\_per_g^{HIV+}(y,p) = 100,000 \sum_{\tau \in year\ y} \left( \frac{\sum\limits_{r \in DR} \sum\limits_{\substack{h \in HIV \\ \{2,3,4\}}} \mu_{6,h,g}(\tau)\ N_{6,r,h,g}(\tau)}{POP_g(\tau,p)} \right) \quad \forall g \in G. \tag{14}$$

Similarly, Equation (15) calculates the number of individuals who died that are HIV negative by gender $g$ at time $\tau$, scaled by population estimates for each gender $g$, summed over all time steps $\tau$ in *year* $y$, and multiplied by 100,000,

$$TBmort\_per_g^{HIV-}(y,p) = 100,000 \sum_{\tau \in year\ y} \left( \frac{\sum\limits_{r \in DR} \mu_{6,1,g}(\tau)\ N_{6,r,1,g}(\tau)}{POP_g(\tau,p)} \right) \qquad \forall g \in G. \tag{15}$$

The aggregated TB mortality rate in *year* $y$, per 100,000 males ($g = 1$) and females ($g = 2$) for program $p$, is equal to the sum of $TBmort\_per_g^{HIV+}(y,p)$ plus $TBmort\_per_g^{HIV-}(y,p)$ by each gender $g$.

#### 4.3 TB and HIV Prevalence

TB prevalence represents the proportion of the population in the model with active TB. HIV prevalence represents the proportion of the population that is living with HIV. We provide TB prevalence by year per 100,000 individuals in Equation (16). Equation (16) calculates the number of individuals with active TB (in TB compartment 6), then sets TB prevalence for *year y* in program $p$ based on the average TB prevalence over time steps $\tau$ in *year y*,

$$TBprev\_per(y,p) = \frac{1}{12} \sum_{\tau \in year\ y} \sum_{r \in DR} \sum_{h \in HIV} \sum_{g \in G} N_{6,r,h,g}(\tau). \tag{16}$$

HIV prevalence represents the proportion of the population who are living with HIV (in HIV compartments 2, 3, and 4). We provide HIV prevalence by gender (per 100,000 males and per 100,000 females) and year in Equation (17). Equation (17) calculates the number of HIV positive individuals at time $\tau$, divided by population estimates for each gender $g$ and multiplied by 100,000, and taking the average over all time steps $\tau$ in *year y*,

$$HIVprev\_per_g(y,p) = 100,000 \left(\frac{1}{12}\right) \sum_{\tau \in year\ y} \left( \frac{\sum_{t \in TB} \sum_{r \in DR} \sum_{\substack{h \in HIV \\ \{2,3,4\}}} N_{t,r,h,g}(\tau)}{POP_g(\tau,p)} \right) \quad \forall g \in G. \tag{17}$$

#### 4.4 Disability-Adjusted Life Years

Disability-adjusted life years (DALYs) are calculated based on years of life lost (YLL) and years lived with disability (YLD) per 100,000 individuals. The years of life lost in *year y* per 100,000 individuals for program $p$, $YLL\_per(y,p)$, is calculated in Equation (18) as a measure that represents the number of years lost from premature mortality for those with active TB and/or HIV positive. Since individuals age out at 60 years old, we assume that any individual who died from HIV/TB would have lived or aged out by the end of the intervention period (in 2028) if they did not die with active TB and/or HIV positive. Equation (18) calculates YLL projected by the model in *year y* for the program $p$, per 100,000 individuals based on the number of individuals departing the active TB compartment (TB compartment 6) and/or HIV positive compartments (HIV 2, 3 and 4). We multiply the number of deaths at time $\tau$ by the time left until the end of the intervention period $(2028 - \tau)$ to represent the number of years each person who died would have lived if they lived to the end of the intervention period,

$$\begin{aligned} YLL\_per(y,p) = & \sum_{\tau \in year\ y} \sum_{r \in DR} \sum_{h \in HIV} \sum_{g \in G} (2028 - \tau)(\mu_{6,h,g}(\tau)\ N_{6,r,h,g}(\tau)) \\ & + \sum_{\tau \in year\ y} \sum_{\substack{t \in TB \\ t \neq 6}} \sum_{r \in DR} \sum_{\substack{h \in HIV \\ \{2,3,4\}}} \sum_{g \in G} (2028 - \tau)(\mu_{t,h,g}(\tau)\ N_{h,r,h,g}(\tau)). \end{aligned} \tag{18}$$

The years lived with disability in *year y* per 100,000 individuals for program $p$, $YLD\_per(y,p)$, is calculated in Equation (19) to quantify the impact on quality of life for those with active TB or living with HIV using disability weights. The parameter $D_{t,h}$ is the disability weight for those in TB compartment $t$ and HIV compartment $h$. Input parameter values for disability weights $D_{t,h}$ are defined for a year living with TB and/or HIV and are given in Section 5.5, Table 7. To convert disability weights to monthly estimates, we multiply disability weights by 1/12,

$$YLD\_per(y,p) = \sum_{\tau \in year\ y} \left( \sum_{t \in TB} \sum_{h \in HIV} \frac{1}{12} D_{t,h} \sum_{r \in DR} \sum_{g \in G} N_{t,r,h,g}(\tau) \right). \tag{19}$$

We calculate disability-adjusted life years (DALYs) for each program $p$ by *year y*, per 100,000 individuals, as the sum of years of life lost (YLL) and years lived with disability (YLD) for each program $p$ by *year y* such that,

$$DALY\_per(y,p) = YLL\_per(y,p) + YLD\_per(y,p). \tag{20}$$

#### 4.5 Program Costs

The cost model includes costs for inpatient and outpatient care for PLWH for causes other than TB, costs to administer TPT, TB treatment costs, and HIV testing costs. The cost in *year y* per 100,000 individuals for program $p$, $Cost\_per(y,p)$, is calculated in Equation (21). These costs are generated by combining HIV prevalence, the number of individuals on TPT, TB prevalence, and ART initiation estimates from model projections with costs associated with each of these states.

We summarise input cost parameters in Table 4. The input cost parameters $Out\_Ocare_h^{COST}$ and $In\_Ocare_h^{COST}$ represents the yearly outpatient and inpatient cost of care for PLWH for causes other than TB in HIV compartment $h$, for $h$ = 2, 3, and 4. The cost to provide a course of TPT is represented by the parameter $IPT^{COST}$. Costs to provide TB treatment are differentiated by drug-resistance status to account for costs associated with the treatment regimens for DS-TB versus MDR-TB, represented by the parameters $TBcare_r^{COST}$, for $r = 1$, and 2. The costs per HIV diagnosis and ART initiation is represented by the parameter $HIVtest^{COST}$.

| Notation | Description |
|---|---|
| | **Costs** |
| $Out\_Ocare_h^{COST}$ | Annual outpatient HIV care for PLWH in HIV compartment $h$, for $h$ = 2, 3, and 4 |
| $In\_Ocare_h^{COST}$ | Annual inpatient HIV care for PLWH in HIV compartment $h$, for $h$ = 2, 3, and 4 |
| $TPT^{COST}$ | Cost to provide a course of TPT |
| $TBcare_r^{COST}$ | Cost to provide a course of TB treatment for those with active TB (in TB compartment 6) in DR compartment $r$, for $r = 1$, and 2 |
| $HIVtest^{COST}$ | Cost of HIV testing to find one person to initiate ART |

Table 4: Description and notation for parameters used in the cost model.

The first line in Equation (21) calculates outpatient and inpatient costs for PLWH (in HIV compartments 2, 3 and 4) for causes other than TB by multiplying the number of individuals in each HIV positive compartment at time $\tau$ by monthly costs of HIV care, where the annual costs of outpatient and inpatient care, $Out\_Ocare_h^{COST}$ and $In\_Ocare_h^{COST}$, are converted to monthly costs by multiplying by 1/12. The second line of Equation (21) calculates the cost to administer TPT by multiplying the number of individuals on TPT at time $\tau$ by monthly costs of administering TPT. Since the TPT cost, $TPT^{COST}$, is defined for an entire course of TPT, we multiply $TPT^{COST}$ by $\omega/12$ to get monthly costs, where $12/\omega$ represents the duration of a TPT course in months. Line three calculates the inpatient and outpatient cost of TB care for those with active TB (in TB compartment 6) and DR compartment $r$ by multiplying the number of individuals with active DS-TB and MDR-TB by monthly TB care costs. The TB care cost, $TBcare_r^{COST}$, is defined for a course of TB treatment. We multiply TB treatment costs by $\upsilon_h \pi_{6,7}/12$ to get monthly costs, where $12/\upsilon_h \pi_{6,7}$ represents the duration of recovery/treatment in months. Line four calculates HIV testing costs by multiplying the number of individuals initiating ART by the average cost per ART diagnosis and ART initiation. To get yearly costs, we sum monthly costs for each time $\tau$ in *year y*,

$$
\begin{aligned}
Cost\_per(y,p) = &\sum_{\tau \in year\ y} \sum_{\substack{h \in HIV \\ \{2,3,4\}}} \frac{1}{12}(Out\_Ocare_h^{COST} + In\_Ocare_h^{COST}) \sum_{t \in TB} \sum_{r \in DR} \sum_{g \in G} N_{t,r,h,g}(\tau) \\
&+ \sum_{\tau \in year\ y} \frac{\omega}{12} IPT^{COST} \sum_{\substack{t \in TB \\ \{2,5\}}} \sum_{r \in DR} \sum_{g \in G} N_{t,r,4,g}(\tau) \\
&+ \sum_{\tau \in year\ y} \sum_{r \in DR} \sum_{h \in HIV} \frac{\upsilon_h \pi_{6,7}}{12} TBcare_r^{COST} \sum_{g \in G} N_{6,r,h,g}(\tau) \\
&+ \sum_{\tau \in year\ y} \sum_{t \in TB} \sum_{r \in DR} \sum_{\substack{h \in HIV \\ \{2,3\}}} \sum_{g \in G} HIVtest^{COST} \eta_{h,4,g}(\tau) N_{t,r,h,g}(\tau). \qquad (21)
\end{aligned}
$$

##### 4.6 Program Health Outcomes, Costs, and Incremental Cost-Effectiveness Ratios

We calculate discounted and undiscounted health outcomes and costs, including TB incident cases, TB deaths, DALYs, and program costs. Undiscounted incident TB cases, TB deaths, DALYs, and costs are accumulated over the 10-year intervention period from 2018 to 2027 for each program $p$, and are specified in Equation (22), Equation (23), Equation (24), and Equation (25), respectively,

$$UTBinc\_total(p) = \sum_{Y \in [2018,2027]} TBinc\_per(y,p) \qquad (22)$$

$$UTBmort\_total(p) = \sum_{Y \in [2018,2027]} TBmort\_per(y,p) \qquad (23)$$

$$UDALY\_total(p) = \sum_{Y \in [2018,2027]} DALY\_per(y,p) \qquad (24)$$

$$UCost\_total(p) = \sum_{Y \in [2018,2027]} Cost\_per(y,p). \qquad (25)$$

Discounted health outcomes and costs are calculated similarly but are multiplied by a discounting factor, $F(y)$. We discount metrics at a rate of 3% or 0.03 to the present value at the start of the intervention period in 2018 for each year in the intervention period, $year\ y \in [2018, 2027]$ as,

$$F(y) = \frac{1}{(1+0.03)^{(y-2018)}}. \qquad (26)$$

Discounted incident TB cases, TB deaths, DALYs, and costs over the 10-year intervention period from 2018 to 2027 are calculated for each program $p$ and converted to 2018 present values with the discounting factor $F(y)$ in Equation (27), Equation (28), Equation (29), and Equation (30), respectively,

$$DTBinc\_total(p) = \sum_{Y \in [2018,2027]} F(y)\ TBinc\_per(y,p) \qquad (27)$$

$$DTBmort\_total(p) = \sum_{Y \in [2018,2027]} F(y)\ TBmort\_per(y,p) \qquad (28)$$

$$DDALY\_total(p) = \sum_{Y \in [2018,2027]} F(y)\ DALY\_per(y,p) \tag{29}$$

$$DCost\_total(p) = \sum_{Y \in [2018,2027]} F(y)\ Cost\_per(y,p). \tag{30}$$

We use these metrics to generate undiscounted and discounted incremental cost-effectiveness ratios (ICERs) to assess the per-dollar cost per incident TB case averted, TB death averted and DALY averted to compare community-based care intervention programs $\tilde{p}$ to programs $p$ including:

1. Program 2 ($\tilde{p} = 2$) to Program 1 ($p = 1$) to evaluate the incremental benefit of a community-based ART intervention,
2. Program 3 ($\tilde{p} = 3$) to Program 1 ($p = 1$) to evaluate the incremental benefit of a community-based ART and TPT intervention and
3. Program 3 ($\tilde{p} = 3$) to Program 2 ($p = 2$) to evaluate the incremental benefit of a nested community-based TPT intervention (assuming community-based ART is already implemented)

We calculate undiscounted ICERs for incident TB cases averted, TB deaths averted, and DALYs averted as in Equation (31), Equation (32), and Equation (33), respectively,

$$UICER^{TBinc}(\tilde{p},p) = \frac{UCost\_total(\tilde{p}) - UCost\_total(p)}{UTBinc\_total(\tilde{p}) - UTBinc\_total(p)} \tag{31}$$

$$UICER^{TBmort}(\tilde{p},p) = \frac{UCost\_total(\tilde{p}) - UCost\_total(p)}{UTBmort\_total(\tilde{p}) - UTBmort\_total(p)} \tag{32}$$

$$UICER^{DALY}(\tilde{p},p) = \frac{UCost\_total(\tilde{p}) - UCost\_total(p)}{UDALY\_total(\tilde{p}) - UDALY\_total(p)}. \tag{33}$$

Discounted ICERs are calculated for incident TB cases averted, TB deaths averted, and DALYs averted as in Equation (34), Equation (35) and Equation (36), respectively,

$$DICER^{TBinc}(\tilde{p},p) = \frac{DCost\_total(\tilde{p}) - DCost\_total(p)}{DTBinc\_total(\tilde{p}) - DTBinc\_total(p)} \tag{34}$$

$$DICER^{TBmort}(\tilde{p},p) = \frac{DCost\_total(\tilde{p}) - DCost\_total(p)}{DTBmort\_total(\tilde{p}) - DTBmort\_total(p)} \tag{35}$$

$$DICER^{DALY}(\tilde{p},p) = \frac{DCost\_total(\tilde{p}) - DCost\_total(p)}{DDALY\_total(\tilde{p}) - DDALY\_total(p)}. \tag{36}$$

### 5 Description of Input Data

This section describes the input data needed to execute the dynamic transmission model and evaluate each program. All the input parameters described in this section can be found at `https://github.com/cgreene3/epi_model_HIV_TB_KZN_SA/tree/master/param_files/input_parameters`. Section 5.1 describes how we initialize the population in the dynamic transmission model at the start of 1940. Section 5.2 describes the input parameter values and calibration ranges used in the dynamic transmission model to describe TB and HIV disease progression in Table 1 and Table 2. In Section 5.3, we describe how we calculate the parameter values for the three care programs, specifically ART coverage and TPT initiation, over the intervention period from the start of 2018 to the end of 2027. Section 5.4 describes how we calibrate the model, Section 5.5 provides disability weights used to calculate DALYs, and finally, Section 5.6 provides the parameter values used in the cost model.

#### 5.1 Population

Initial population state values at the start of 1940 are assigned according to estimates for 1990 on the total population, TB prevalence, and LTBI prevalence by gender from GBD 2019 for 15 to 59-year-olds from KwaZulu-Natal, South Africa [4]. We scale the initial population state values proportionally to 100,000 to match our model population. The non-zero population compartments include TB compartments 1 (Uninfected, not on TPT), 3 (LTBI, recent), 4 (LTBI, remote), and 6 (active TB), HIV compartment 1 (HIV-), and both gender compartments. GBD 2019 estimates the total population, active TB and LTBI for 1990 by gender. To estimate the number of individuals uninfected (in TB compartment 1) we subtract TB prevalence and LTBI prevalence estimates from total population estimates. The initial population state values can be found at `https://github.com/cgreene3/epi_model_HIV_TB_KZN_SA/tree/master/param_files/input_parameters/pop_init_df_1940.csv`.

#### 5.2 Parameter Values that Describe TB and HIV Disease Progression

Table 5 includes the input parameter values and calibration ranges for the parameters used in the dynamic transmission model in Table 1 and Table 2. We used a range of 25% above and below the mean value from cited sources for calibrated parameters. We calibrate a total of 34 parameters. Most input parameters are not time-dependent. Time-varying input parameters are notated as functions of $\tau$ in Table 5 and include rate of TPT initiation for HIV compartment $h$ and gender $g$, denoted $\kappa_{h,g}(\tau)$ as described in Section 5.2.1, HIV incidence rates for gender $g$, denoted $\eta_{1,2,g}(\tau)$ as described in Section 5.2.2, ART coverage for gender $g$, denoted $\sigma_g(\tau)$ as described in Section 5.2.3, the proportion of PLWH (not on ART, with CD4 > 200) of gender $g$ eligible to initiate ART, denoted $\varrho_g(\tau)$ as described in Section 5.2.4, mortality rates for TB compartment $t$, HIV compartment $h$ and gender $g$, denoted $\mu_{t,h,g}(\tau)$ as described in Section 5.2.5, and allocation of births to TB compartment $t$, DR compartment $r$, HIV compartment $h$ and gender compartment $g$, denoted $\alpha^{in}_{t,r,h,g}(\tau)$ as described in Section 5.2.6.

Table 5: Input parameters used in the dynamic transmission model to describe TB and HIV disease progression. The mean value for the 34 calibrated parameters is given, with the 25% calibration range used in calibration. Time-varying parameters, notated as a function of $\tau$, are described in Section 5.2.1 to Section 5.2.6. care program-specific parameters are highlighted in green. HIV infection rates are highlighted in blue to indicate that they are indirectly impacted by program.

| Parameter description | Value [Calibration Range] | Reference |
|---|---|---|
| **Parameters that Impact TB Force of Infection** | | |
| $\beta_g$ Number of effective contacts for TB transmission per year for: | | [46] |
| Males, $\beta_1$ | 14 [10.5, 17.5] | |
| Females, $\beta_2$ | 14 [10.5, 17.5] | |
| $\phi_h$ Relative transmissibility of TB for those: | | [21,30] |
| HIV−, $\phi_1$ | 1 (fixed value) | |
| HIV+, Not on ART, CV4 > 200, $\phi_2$ | 0.9 [0.675, 1.125] | |
| HIV+, Not on ART, CV4 $\leq$ 200, $\phi_3$ | 0.6 [0.45, 0.75] | |
| HIV+, On ART, $\phi_4$ | 0.9 [0.675, 1.125] | |
| Continued on next page | | |

**Table 5 – continued from previous page**

| Parameter description | Value [Calibration Range] | Reference |
|---|---|---|
| $\varepsilon$ Fraction of new TB infections that are MDR-TB | 0.037 [0.02775, 0.04625] | [50] |
| $\iota_r$ Diminished risk of acquiring a latent TB infection of strain $r$ while uninfected and on TPT, such that: | | |
| DS-TB, $\iota_1$ | 0.43 [0.3225, 0.5375] | [14] |
| MDR-TB, $\iota_2$ | 1 (fixed value) | |
| $\xi$ Increased risk of reinfection after recovering/treatment of active TB | 4 [3, 5] | [47] |
| $\zeta$ Partially protective effect of LTBI against acquiring a new TB infection | 0.4 [0.3, 0.5] | [14,21,27,47] |
| **Parameters that Describe TB Progression** | | |
| $1/\omega$ Duration of TPT course (years) | 0.5 (fixed value) | [7] |
| $\gamma_r$ Indicator to reflect that those infected with MDR-TB do not yield benefits of TPT, such that: | | |
| Drug-Susceptible strain, $\gamma_1$ | 1 (fixed value) | |
| MDR strain, $\gamma_2$ | 0 (fixed value) | |
| $1/\pi_{3,4}$ Duration of LTBI recently infected period (years) | 2 (fixed value) | [48] |
| $\pi_{i,6}$ Rates of TB progression from: | | |
| Recent LTBI to active TB, $\pi_{3,6}$ | 0.025 [0.01875, 0.03125] | [15,29] |
| Remote LTBI to active TB, $\pi_{4,6}$ | 0.001 [0.00075, 0.00125] | [12,17] |
| LTBI on TPT to active TB, $\pi_{5,6}$ | 0.0113 [0.0085, 0.0142] | [38] |
| Recovered/Treated to active TB, $\pi_{7,6}$ | 0.01 [0.0075, 0.0125] | [11] |
| LTBI after TPT to active TB, $\pi_{8,6}$ | 0.0113 [0.0085, 0.0142] | [38] |
| $\theta_h$ Relative risk of TB progression for individuals that are: | | |
| HIV−, $\theta_1$ | 1 (fixed value) | |
| HIV+, not on ART, CD4 $>$ 200, $\theta_2$ | 10 [7.5, 12.5] | [6,49] |
| HIV+, not on ART, CD4 $\leq$ 200, $\theta_3$ | 17 [12.75, 21.25] | [2,9] |
| HIV+, on ART, $\theta_4$ | 3 [2.25, 5.25] | [26,45] |
| $1/\pi_{6,7}$ Duration of treatment/recovery (years) | 0.5 (fixed value) | [24] |
| $1/(\upsilon_h\pi_{6,7})$ Duration of active TB (years) that reflect delays in treatment for: | | [24] |
| HIV−, $1/(\upsilon_1\pi_{6,7})$ | 2 [1.6, 2.7] | |
| HIV+, not on ART, CD4 $>$ 200, $1/(\upsilon_2\pi_{6,7})$ | 1.5 [1.2, 2] | |
| HIV+, not on ART, CD4 $\leq$ 200, $1/(\upsilon_3\pi_{6,7})$ | 1.5 [1.2, 2] | |
| HIV+, on ART, $1/(\upsilon_4\pi_{6,7})$ | 1.5 [1.2, 2] | |
| $\kappa_{h,g}(\tau)$ Rate of TPT initiation for HIV compartment $h$ and gender $g$ at time $\tau$. | Time-varying (fixed value) | [7] |
| **Parameters that Describe HIV Progression** | | |
| | | Continued on next page |

**Table 5 – continued from previous page**

| Parameter description | Value [Calibration Range] | Reference |
|---|---|---|
| $\eta_{1,2,g}(\tau)$ HIV incidence time-varying rates for gender $g$ at time $\tau$ calculated based on mean HIV incidence rates for *year* $y$, $\eta_{1,2,g}^{VAL}(y)$ and the calibrated factor $\eta_{1,2,g}^{FACTOR}$ for: | Time-varying (calibrated) | [4, 37] |
| Males $\eta_{1,2,1}^{FACTOR}$ | 1 [0.75, 1.25] | |
| Females $\eta_{1,2,2}^{FACTOR}$ | 1 [0.75, 1.25] | |
| $1/\eta_{2,3,g}$ Duration from HIV acquisition to CD4 $< 200$ (years) for: | | [39] |
| Males $1/\eta_{2,3,1}$ | 7.72 [5.79, 9.65] | |
| Females $1/\eta_{2,3,2}$ | 10.25 [7.6875, 12.8125] | |
| $\sigma_g(\tau)$ ART coverage for gender $g$ at time $\tau$ | Time-varying (fixed value) | [7, 41] |
| $\varrho_g(\tau)$ Proportion of gender $g$ eligible to initiate ART at time $\tau$. | Time-varying (fixed value) | [37] |
| **Parameters that Describe Morality Rates** | | |
| $\mu_{t,h,g}(\tau)$ Mortality rates time-varying rates for those in TB compartment $t$, HIV compartment $h$, and gender compartment $g$ calculated based on yearly mean baseline mortality rates, $\mu_g^{VAL}(y)$, calibrated increase risk of disease mortality $R_{t,h}$ (see below), and the calibrated factor $\mu_g^{FACTOR}$ for: | Time-varying (calibrated) | [4] |
| Males $\mu_1^{FACTOR}$ | 1 [0.75, 1.25] | |
| Females $\mu_2^{FACTOR}$ | 1 [0.75, 1.25] | |
| $R_{t,h}$ Increased risk of mortality for those in TB compartment $t$ and HIV compartment $h$: | | [2, 6, 34–36] |
| No active TB and HIV-, $R_{t,1}, t \neq 6$ | 1 (fixed value) | |
| No active TB and HIV+, not on ART, CD4 $> 200$, $R_{t,2}, t \neq 6$ | 8 [6, 10] | |
| No active TB and HIV+, not on ART, CD4 $\leq 200$, $R_{t,3}, t \neq 6$ | 26 [19.5, 32.5] | |
| No active TB and HIV+, on ART, $R_{t,4}, t \neq 6$ | 1.35 [1.2, 1.5] | |
| Active TB and HIV-, $R_{6,1}$ | 15.5 [11.625, 19.375] | |
| Active TB and HIV+, not on ART, CD4 $> 200$, $R_{6,2}$ | 26 [19.5, 32.5] | |
| Active TB and HIV+, not on ART, CD4 $\leq 200$, $R_{6,3}$ | 50 [37.5, 62.5] | |
| Active TB and HIV+, on ART, $R_{6,4}$ | 18.5 [13.875, 23.125] | |
| **Parameters that Describe Entries and Exits due to Aging** | | |
| $\alpha_{t,r,h,g}^{in}(\tau)$ Proportion of the population that enters into TB compartment $t$, DR compartment $r$, HIV compartment $h$, and gender compartment $g$ and time $\tau$ | Time-varying (fixed value) | [4, 50] |
| $\alpha^{ageout}$ Rate of exit from the population due to aging | $\frac{1}{60-15}$ | |

##### 5.2.1 TPT initiation rates

We only allow those in HIV compartment 4 (HIV+, on ART) to initiate TPT, so TPT initiation rates are set to zero for those not in HIV compartment 4, i.e., $\kappa_{1,g}(\tau) = \kappa_{2,g}(\tau) = \kappa_{3,g}(\tau) = 0$ for all $\tau$. For those in HIV compartment 4, TPT initiation rates $\kappa_{4,g}(\tau)$ change over time according to TPT availability and program (during the intervention period). From the start of 1940 to the end of 2004, TPT initiation rates are set to zero for those in HIV compartment 4, i.e., $\kappa_{4,g}(\tau) = 0$, to indicate that TPT was not widely available. TPT initiation rates are linearly interpolated from zero in 2004 (the year before TPT became widely available) to the value in 2018 for the standard facility-based ART and TPT program (Program 1). During the intervention period, TPT initiation rate $\kappa_{4,g}(\tau)$ is held constant from the start of 2018 to the end of 2027, and is differentiated by program as given in Table 6 and discussed in Section 5.3.2. The yearly TPT initiation rate values by program are available at `https://github.com/cgreene3/epi_model_HIV_TB_KZN_SA/blob/master/param_files/input_parameters/ipt_initiation_df.csv`.

##### 5.2.2 HIV Incidence Rates

HIV incidence rates, $\eta_{1,2,g}(\tau)$ for gender $g$, vary over time according to mean yearly HIV incidence rate estimates $\eta^{VAL}_{1,2,g}(y)$. A multiplication factor, $\eta^{FACTOR}_{1,2,g}$, is used to adjust all yearly HIV incidence rate estimates, and is calibrated as indicated in Table 5. We multiply each mean yearly incidence rate estimate by the multiplication factor, and we assume the monthly incidence rate is the same for all months within the year, that is,

$$\eta_{1,2,g}(\tau) = \eta^{FACTOR}_{1,2,g}\eta^{VAL}_{1,2,g}(y) \quad \text{for} \quad g \in G,\ \tau \in year\ y.$$

From 1940 to 1979, the mean yearly HIV incidence estimates are set to zero, i.e., $\eta^{VAL}_{1,2,g}(y) = 0$, to represent that we assume that is no HIV incidence in the model. We introduce HIV incidence at the beginning of 1980. Between 1980 and 1990, the mean yearly HIV incidence estimates are linearly interpolated from zero in 1979 to the value in 1990. Between 1990 to 2017, the mean yearly HIV incidence estimates are set to yearly estimates from GBD 2019 for males and females between the ages of 15 to 59-year-olds in KwaZulu-Natal, South Africa [4]. For the years in the intervention period, starting at the beginning of 2018 and continuing through the end of 2027, the mean yearly HIV incidence estimates are projected for each program by applying rate of changes, denoted $\eta^{RATEOFCHANGE}_{1,2,g}(y)$ for $year\ y$, that decrease HIV incidence estimates from the prior year, as

$$\eta^{VAL}_{1,2,g}(y) = \eta^{RATEOFCHANGE}_{1,2,g}(y)\eta^{VAL}_{1,2,g}(y-1).$$

The rate of changes $\eta^{RATEOFCHANGE}_{1,2,g}(y)$, are projected by the Data-driven Recommendations for Interventions against Viral Infection (DRIVE) model [37] using ART coverage values for each program provided in Table 6. While the DRIVE model [37] projects that HIV incidence will decrease each year for all programs, it projects that HIV incidence for community-based ART programs (Program 2 and 3) will decrease at a higher rate than the facility-based ART program (Program 1) due to the indirect impacts of increased ART coverage on HIV incidence rates.

HIV incidence yearly estimates for males $\eta^{VAL}_{1,2,1}(y)$ and females $\eta^{VAL}_{1,2,2}(y)$ for each program are represented in Figure 3, where the values for Programs 2 and 3 are differentiated from Program 1 during the intervention period for the years between 2018 and 2027. The mean yearly HIV estimates between 1990 to 2017 from GBD 2019 can be found at github/epi_model_HIV_TB_KZN_SA/param_files/calculated_param_gen/raw_input_data/GBD/hiv_inc_num.csv, the rate of change projections, $\eta_{1,2,g}^{RATEOFCHANGE}(y)$ can be found at github/epi_model_HIV_TB_KZN_SA/param_files/calculated_param_gen/raw_input_data/DO_ART/intervention_hiv_incidence_est.xlsx and the calculated HIV incidence yearly mean estimate values can be found at https://github.com/cgreene3/epi_model_HIV_TB_KZN_SA/blob/master/param_files/input_parameters/hiv_inc_df.csv.

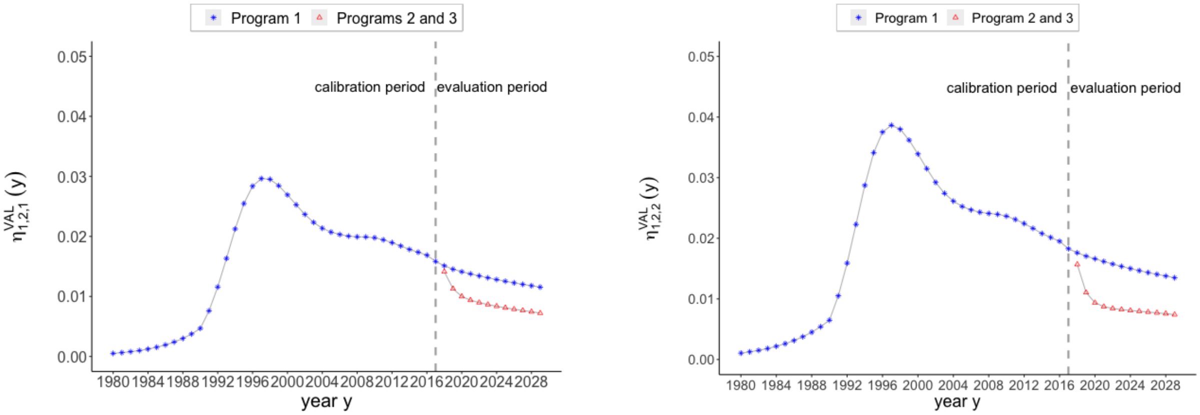

(a) Mean HIV incidence rate, for *year* $y$ and males, $\eta_{1,2,1}^{VAL}(y)$

(b) Mean HIV incidence rate, for *year* $y$ and females, $\eta_{1,2,2}^{VAL}(y)$

Figure 3: Yearly mean HIV incidence estimates for males, $\eta_{1,2,1}^{VAL}(y)$, and females, $\eta_{1,2,2}^{VAL}(y)$ from 1980 to 2028. Over the intervention period between 2018 and 2027, HIV estimates are differentiated by program. HIV incidence rate estimates for Program 1 are in blue (stars), and incidence rate estimates for Programs 2 and 3 are in red (triangles).

##### 5.2.3 ART Coverage

ART coverage $\sigma_g(\tau)$ changes over time according to ART availability in KwaZulu-Natal, South Africa, and program as described in Section 2.1.2. Before 2004, ART is not available, so ART coverage for both genders are set to zero, i.e., $\sigma_1(\tau) = \sigma_2(\tau) = 0$, to represent that ART is not available until the start of 2004. Between 2004 (when ART becomes available) and 2018, ART coverage estimates are linearly interpolated from zero in 2003 to the value in 2018 for the standard facility-based ART and TPT program (Program 1). During the intervention period, ART coverage is differentiated by program and held constant from the beginning of 2018 to the end of 2027, with the values for $\sigma_g(\tau)$ provided in Table 6 and discussed in Section 5.3.1. The yearly ART coverage input values by program are available at https://github.com/cgreene3/epi_model_HIV_TB_KZN_SA/blob/master/param_files/input_parameters/art_coverage_df.csv.

##### 5.2.4 Proportion of PLWH (not on ART, with CD4 >200) Eligible to Initiate ART

The proportion of PLWH not on ART with CD4 >200 that are eligible to initiate ART, denoted $\varrho_g(\tau)$ by gender $g$ at time $\tau$, correspond to changes in clinical guidance on ART eligibility described in Section 2.1.2. Between the start of 1940 and the end of 2010, no one in HIV compartment 2 (HIV+, not on ART, CD4 > 200) is eligible to initiate ART, i.e., $\varrho_g(\tau) = 0$. Between the start of 2011 to the end of 2015, the proportions of those in HIV compartment 2 (HIV+, not on ART, CD4 > 200) by gender that are eligible to initiate ART are set to estimates projected by the DRIVE model [37], such that $0 < \varrho_g(\tau) < 1$. From the start of 2016, all those in HIV compartment 2 (HIV+, not on ART, CD4 > 200) are eligible to initiate ART, i.e., $\varrho_g(\tau) = 1$. The input parameter values are available at `https://github.com/cgreene3/epi_model_HIV_TB_KZN_SA/blob/master/param_files/input_parameters/art_prop_eligible_df.csv`.

##### 5.2.5 Mortality Rates by TB and HIV Compartments

Mortality rates for populations in TB compartment $t$, HIV compartment $h$ for gender $g$ at time $\tau$, denoted $\mu_{t,h,g}(\tau)$, represent mortality due to causes that are not TB-related or HIV-related and vary over time according to yearly mean baseline mortality rate estimates, denoted $\mu_g^{VAL}(y)$, a multiplication factor used to adjust all yearly mean baseline mortality rate estimates, denoted $\mu_g^{FACTOR}$, and an increased risk of mortality for those with TB or HIV, denoted $R_{t,h}$. The increased risk of mortality for those in TB compartment $t$ and HIV compartment $h$, $R_{t,h}$, and the multiplication factor by gender, $\mu_g^{FACTOR}$, is calibrated, with the ranges for calibration provided in Table 5. To obtain $\mu_{t,h,g}(\tau)$, we multiply the mean baseline mortality rates for each gender by the increased risk of mortality and the multiplication factor, and we assume the monthly mortality rate is the same for all months within the year, that is,

$$\mu_{t,h,g}(\tau) = \mu_g^{FACTOR}\ R_{t,h}\ \mu_g^{VAL}(y) \quad \text{for} \quad t \in TB, h \in HIV, g \in G,\ \tau \in year\ y.$$

The yearly baseline mortality rates, $\mu_g^{VAL}(y)$, are calculated in Equation (37) using yearly GBD 2019 estimates (provided for the years 1990 to 2017) on the number of deaths from all causes denoted $GBD_g^{ALLMORT}(y)$, the number of TB-related deaths denoted $GBD_g^{TBMORT}(y)$, the number of HIV-related deaths not related to TB denoted $GBD_g^{OHIVMORT}(y)$, and population estimates denoted $GBD_g^{POP}(y)$ by gender $g$ in *year* $y$ for males and females between the ages of 15 to 59-year-olds in KwaZulu-Natal, South Africa. In Equation (37), we subtract out the number of TB-related and HIV-related deaths from all causes to calculate the number of deaths from causes not related to TB or HIV and divide by population estimates to convert the number of deaths into a rate. We use this data to calculate the yearly baseline mortality rates, $\mu_g^{VAL}(y)$, for the years 1990 to 2017 as,

$$\mu_g^{VAL}(y) = \frac{GBD_g^{ALLMORT}(y) - GBD^{TBMORT}(y) - GBD^{OHIVMORT}(y)}{GBD_g^{POP}(y)}. \tag{37}$$

From the start of 1940 to the end of 1989, the yearly baseline mortality rates, $\mu_g^{VAL}(y)$ are set to the 1990 estimate $\mu_g^{VAL}(y)$, with year $y = 1990$. During the intervention period, for the years 2018 to 2027, yearly baseline mortality rates are held constant at 2017 estimates. Mean yearly baseline mortality rates $\mu_g^{VAL}(y)$ for each gender $g$ for *year* $y$ are illustrated in Figure 4. The data for all-cause mortality, $GBD_g^{ALLMORT}(y)$, TB-related mortality, $GBD_g^{TBMORT}(y)$, HIV-related mortality for causes other than TB, $GBD_g^{OHIVMORT}(y)$, and population estimates denoted $GBD_g^{POP}(y)$ are available at `https://github.com/cgreene3/epi_model_HIV_TB/south_africa/param_files/calculated_param_gen/raw_input_data/GBD`. The calculated baseline mortality estimate values can be found at `https://github.com/cgreene3/epi_model_HIV_TB/south_africa/param_files/baseline_mortality.csv`.

##### 5.2.6 Allocation of Births to TB and HIV Compartments

A proportion of births entering the model at time $\tau$, $\alpha_{t,r,h,g}^{in}(\tau)$, are based on GBD 2019 projections for 10 to 15-year-olds from KwaZulu-Natal, South Africa [4]. GBD 2019 provides projections

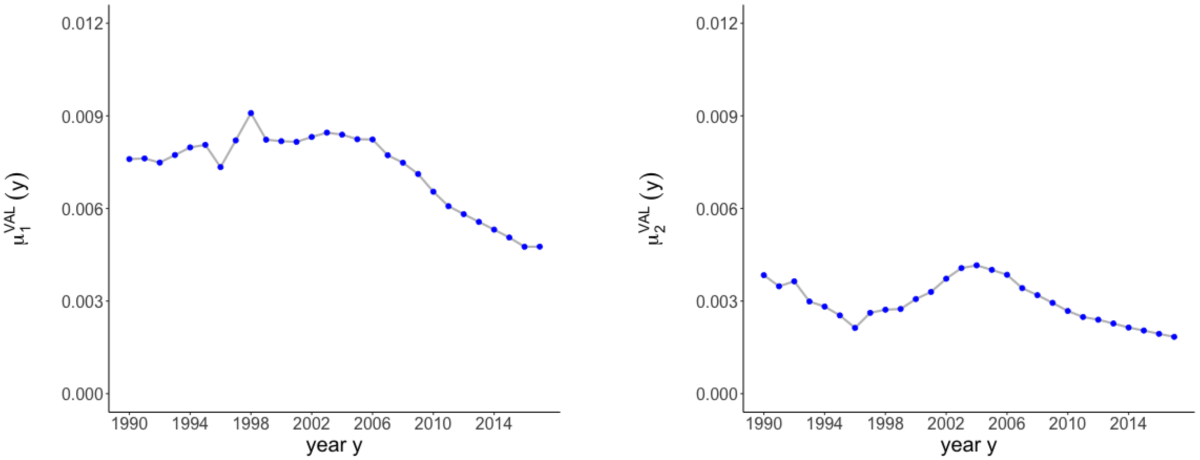

(a) Mean baseline mortality rates, for *year y* and males, $\mu_1^{VAL}(y)$

(b) Mean baseline mortality rates, for *year y* and females, $\mu_2^{VAL}(y)$

Figure 4: Mean yearly baseline mortality rate estimates for males, $\mu_1^{VAL}(y)$, and females, $\mu_2^{VAL}(y)$ from 1990 to 2017 [4].

on the number of males and females with active TB, living with HIV, LTBI, and population estimates for the years 1990 to 2017. We use these estimates to allocate a proportion of births to TB compartments 1 (Uninfected, not on TPT), 3 (LTBI, recent), 4 (LTBI, remote), and 6 (active TB), distinguished by drug resistance, HIV status, and gender that are representative of changes in population characteristics over time. We scale population estimates in each of these compartments for each year to percentages so that the total number of individuals aging into the model equals the total number of individuals aging out of the model as described in Section 2.1.3.

Since GBD 2019 does not estimate the number of individuals uninfected with TB (in TB compartment 1) and HIV negative, we calculate the number of males and females uninfected with TB (TB compartment 1) and HIV negative (HIV compartment 1) by subtracting out the number of males and females with active TB, LTBI or HIV positive from population estimates. We do not allocate births to TB compartments associated with TPT (TB compartments 2, 5 and 8) or recovery from TB (TB compartment 7), such that $\alpha^{in}_{t,r,h,g}(\tau) = 0$ for $t = \{2, 5, 7, 8\}$ for all $\tau$.

GBD 2019 does not distinguish LTBIs by LTBI infected recently (TB compartment 3) and LTBI infected remotely (TB compartment 4). We assume that those aging into the model (on their 15th birthday) were equally likely to be exposed in each year prior to aging into the model, such that 2/15 of LTBIs are recent (exposed within the past two years), and 13/15 of LTBI are remote (exposed more than two years ago). GBD 2019 study also does not differentiate LTBI projections by drug resistance or HIV status. We assume that the proportion of LTBIs that are MDR corresponds to estimates from the 2020 Global Tuberculosis Report [2] and that the proportion of LTBIs that are associated with PLWH correspond to yearly HIV prevalence estimates from GBD 2019. GBD 2019 does not provide HIV prevalence among levels of HIV disease severity, so we assign all PLWH who age into the model into HIV compartment 2 (HIV+, not on ART, CD4 > 200).

For $\tau$ associated with the years between 1940 and 1979 (before HIV incidence is introduced in the model) new entries are only assigned to HIV compartment 1 (HIV-) according to 1990 estimates (aggregated over HIV status), such that $\alpha^{in}_{t,r,h,g}(\tau) = 0$ for $h = \{2, 3, 4\}$ for all time steps $\tau$ associated with the years 1940 to 1979. For $\tau$ associated with the years between 1980 and 1989, $\alpha^{in}_{t,r,h,g}(\tau)$ is set to 1990 estimates. Between the start of 1990 to the end of 2017 the proportion of births allocated to each compartment changes according to yearly estimates. From 2018 and 2027, we assign new entries based on 2017 estimates. Yearly estimates for the total population, TB prevalence, and LTBI prevalence by gender and HIV status as projected by GBD 2019 for 10 to 15-year-olds from KwaZulu-Natal, South Africa [4] can be found at `https://github.com/cgreene3/epi_model_HIV_TB/south_africa/param_files/calculated_param_gen/raw_input_data/GBD`. The calculated aging in proportions can be found at `https://github.com/cgreene3/epi_model_HIV_TB_KZN_SA/blob/master/param_files/input_parameters/birth_perc_df_overtime.csv`.

##### 5.3 Parameter Values for Care Programs

We model three care programs, including standard facility-based ART and TPT care ($p = 1$), community-based ART with standard facility-based TPT care ($p = 2$), and community-based ART with TPT care ($p = 3$) from the beginning of 2018 to the end of 2027. The input parameters that depend on the care program include ART coverage for gender $g$, denoted $\sigma_g(\tau)$, and TPT initiation rates for those in HIV compartment $h$ and gender $g$, $\kappa_{h,g}(\tau)$. ART coverage for gender $g$, $\sigma_g(\tau)$, represents the proportion of PLWH on ART, that is, the ratio of those in HIV compartment 4 to those in HIV positive compartments (in HIV compartments 2, 3, and 4). TPT initiation rates, $\kappa_{h,g}(\tau)$, represent the proportion of those in HIV compartment $h$ and gender $g$ that initiate TPT. We assume only PLWH on ART (in HIV compartment 4) initiate TPT, such that $\kappa_{h,g}(\tau) = 0,\ h = \{1, 2, 3\}$.

ART coverage and TPT initiation rates vary for each program based on observations from the DO ART trial [7]. The impact of the three programs on ART coverage and TPT initiation rates among PLWH on ART over the program intervention period from the beginning of 2018 to the end of 2027 are provided in Table 6. These input parameters are held constant over the intervention period for the times associated with the years between 2018 and 2027.

Program 1, standard facility-based ART and TPT care, reflects the levels of ART coverage and TPT initiation rates among PLWH on ART observed in the facility-based care arm of the DO ART trial [7]. Program 3, community-based ART with TPT care, reflects the levels of ART coverage and TPT initiation rates observed in the community-based care arm of the DO ART trial. Community-based ART was only tested in the DO ART trial with a nested community-based TPT care program (Program 3); however, we additionally test community-based ART without community-based TPT (Program 2) to test the independent effects of community-based ART on TB incidence and TB mortality. We do this by setting ART coverage for Program 2 to the same values as in Program 3. We back out the TPT initiation percentage among PLWH on ART for Program 2 by equating the TPT initiation percentage among all PLWH (including those on ART and not on ART) to the values from Program 1.

###### 5.3.1 ART Coverage

ART coverage for gender $g$, $\sigma_g(\tau)$, varies for each program, based on the proportion of participants in the DO ART trial that achieved viral suppression, denoted $VS_g$, and accounts for the proportion of PLWH on ART of gender $g$ who achieve viral suppression with the parameter $AT_g$, and the proportion of PLWH diagnosed with HIV by gender $g$ with the parameter $DH_g$ in Equation (38). The proportion of PLWH on ART of gender $g$ who achieve viral suppression, $AT_g$, and the proportion of PLWH diagnosed with HIV of gender $g$, $DH_g$, are based on findings from the 2017 South African National HIV Prevalence, Incidence, Behaviour, and Communication Survey [41]. We assume all those who are virally suppressed are on ART; however, some proportion of those on ART are not virally suppressed. We divide the proportions of male and female participants

| **Program Number** ($p$) **and Name** | **ART Coverage %** $\sigma_g(\tau)$ | | **TPT initiation % among PLWH on ART** $\kappa_{h,g}(\tau), h=4$ | |
|---|---|---|---|---|
| | *Male* $g=1$ | *Female* $g=2$ | *Male* $g=1$ | *Female* $g=2$ |
| **1. Standard facility-based ART and TPT care** ($p=1$) | 49 | 69 | 29 | 27 |
| **2. Community-based ART with standard facility-based TPT care** ($p=2$) | 82 | 83 | 17 | 22 |
| **3. Community-based ART with TPT care** ($p=3$) | 82 | 83 | 70 | 75 |

Table 6: Care programs and parameter values for time periods associated with the years in the intervention period from 2018 to 2027.

that achieved viral suppression in the DO ART trial, $VS_g$, by the proportion of PLWH on ART of gender $g$ who achieve viral suppression, $AT_g$, in Equation (38) to calculate the proportion of participants in the trial who are on ART, including participants that are both virally suppressed on ART and not virally suppressed on ART. All participants in the trial on ART know their status; however, our model considers PLWH who do not know their status. We calculate the proportion of the HIV positive population on ART by gender $g$, $\sigma_g(\tau)$, in Equation (38) by multiplying the proportion of participants in the trial who are on ART ($VS_g/AT_g$) by the proportion of PLWH diagnosed with HIV, $DH_g$, as

$$\sigma_g(\tau) = DH_g \left(\frac{VS_g}{AT_g}\right) \qquad \forall g \in G. \tag{38}$$

Under standard facility-based ART and TPT care (Program 1) in the DO ART trial, 51% of male participants and 70% of female participants achieved viral suppression, i.e., $VS_1 = 51\%, VS_2 = 70\%$. Under community-based ART with TPT care (Program 3) in the DO ART trial, 72% of male participants and 73% of female participants achieved viral suppression, i.e., $VS_1 = 72\%, VS_2 = 73\%$ [4].

The 2017 South African National HIV Prevalence, Incidence, Behaviour, and Communication Survey [41] estimates 82% of males, 90% of females, and 88% of both males and females are virally suppressed given they are on ART. Under standard facility-based ART and TPT care (Program 1), we set $AT_1 = 82\%$ and $AT_2 = 90\%$. Under community-based ART with TPT care (Program 3), we set the proportion of PLWH who achieve viral suppression to be the same for both genders, $AT_1 = AT_2 = 88\%$.

The 2017 South African National HIV Prevalence, Incidence, Behaviour, and Communication Survey [41] estimates 78% and 90% of males and females living with HIV know their status. Under Program 1, we set $DH_1 = 78\%, DH_2 = 90\%$. The DO ART trial includes community-wide HIV testing, so under the community-based ART with TPT care program (Program 3), we assume that all PLWH know their status, so we set $DH_1 = DH_2 = 100\%$.

These values are used to generate the ART coverage estimates as presented in Table 6 for Program 1 for males ($78\% \times \frac{51\%}{82\%} = 49\%$) and females ($90\% \times \frac{70\%}{90\%} = 69\%$); and Program 3 for males ($100\% \times \frac{72\%}{88\%} = 82\%$) and females ($100\% \times \frac{73\%}{88\%} = 83\%$). We assume the same ART coverage under Program 2 as in Program 3.

##### 5.3.2 TPT Initiation Rates

TPT initiation rates $\kappa_{h,g}(\tau)$, $h = 4$ for Program 1 (standard facility-based ART and TPT care) are based on observations from the DO ART trial of the proportion of PLWH on ART who initiated TPT in the standard facility-based ART and TPT care trial group (29% among men on ART, 27% among women on ART). TPT initiation rates for Program 3 are also based on observations from the DO ART trial of the proportion of PLWH on ART who initiated TPT in the community-based ART and TPT care group (70% among men on ART, 75% among women on ART).

Program 2 was not observed in the DO ART trial but is modeled in the current study to test the independent effect of community-based ART without changing the proportion of PLWH who initiate TPT. We infer the proportion of PLWH on ART who initiate TPT by gender $g$, $\kappa_{h,g}(\tau), h = 4$ for Program 2, so that TPT initiation rates among all PLWH are the same in Program 1 as in Program 2, while accounting for the differences in ART coverage under community-based ART care. The TPT initiation rates among all PLWH for Program 1 is approximately equal to $\sigma_g \kappa_{4,g}(\tau)$, i.e., for males $40\% \times 29\% = 0.1421$, and for females $69\% \times 27\% = 0.1863$. Given values for $\sigma_g(\tau)$ for Program 2, as in Table 6, we solve for $\kappa_{4,g}(\tau)$ for Program 2. Hence, for Program 2, for males, $\kappa_{4,1}(\tau) = \frac{0.1421}{82\%} = 17\%$, and for females, $\kappa_{4,2}(\tau) = \frac{0.1863}{83\%} = 22\%$.

#### 5.4 Calibration

We calibrate the model over 34 parameters shown in Table 5. We use Latin hypercube sampling to generate 100,000 parameter sets from the ranges specified in Table 5 for all 34 calibrated parameters [44]. The 100,000 parameter sets can be found at `https://github.com/cgreene3/epi_model_HIV_TB_KZN_SA/blob/master/param_files/input_parameters/calibration_sets_df.csv`. We run the model for each parameter set under the standard facility-based ART and TPT care program (Program 1) from the start of 1940 to the end of 2017. We consider ten metrics for calibration, including TB incidence rates by HIV status (HIV- and HIV+) and gender, TB mortality rates by HIV status (HIV- and HIV+) and gender, and HIV prevalence by gender. These metrics are calculated from model outputs for each of the 100,000 parameter sets using the equations provided in Section 4. We evaluate ten metrics in 2005, the peak of the HIV epidemic, and in 2017, the year prior to the start of the intervention period of the DO ART trial. We compare model values to corresponding target calibration ranges in 2005 and 2017 based on 95% uncertainty intervals from the GBD Study 2019 for adults between 15 and 59 years old in KwaZulu-Natal, South Africa [4]. The target calibration ranges can be found at `https://github.com/cgreene3/epi_model_HIV_TB_KZN_SA/tree/master/param_files/target_calibration_estimates`. We accept a parameter set if all 20 model values fall into all 20 target calibration ranges and shows a reduction in the total number of TB cases and deaths over the intervention period in Program 3 versus Program 2. The calibration results in 859 accepted parameter sets. The accepted parameter sets can be found at `https://github.com/cgreene3/epi_model_HIV_TB_KZN_SA/blob/master/calibration_analysis/accepted_calibration_sets_ref_df.csv`. Figure 5 illustrates the mean, minimum and maximum model values of the 859 accepted parameter sets for each of the ten calibration metrics between the years 1990 to 2017. In 2005 and 2017, when we compare model values to target calibration ranges, target calibration ranges are emphasized lines. Figure 6 illustrates the distribution of accepted parameter sets for rates of TB progression from recent LTBI (TB compartment 3), remote LTBI (TB compartment 4), LTBI on TPT (TB compartment 5), and LTBI after TPT (TB compartment 8) to active TB (TB compartment 6). The distributions of accepted parameters for all 34 calibrated parameters can be found at `https://github.com/cgreene3/dynamic_transmission_model_HIV_TB_KZN_SA/tree/master/param_files/distribution_of_accepted_` points_graphs.

#### 5.5 Parameter Values for Disability-Adjusted Life Years

We use disability weights in conjunction with projected model states to calculate Disability-Adjusted Life Years (DALYs) as described in Section 4.4. Disability weights for those in TB compartment $t$ and HIV compartment $h$, denoted $D_{t,h}$ are provided in Table 7. Those without active TB (not in TB compartment 6) or living with HIV (in TB compartment 1) have an associated disability weight of zero. Disability weights associated with active TB (in TB compartment 6) or HIV (in HIV compartments 2, 3, and 4) are from GBD 2019 [16].

The disability weights associated with PLWH without active TB from GBD 2019 include stratification over CD4 levels, ART status and anemia levels. Since we do not track anemia levels of PLWH in the dynamic transmission model, we apply the disability weight corresponding to mild anemia to all PLWH (in HIV compartments 2, 3 and 4) without active TB (not in TB compartment 6) since mild anemia is common among PLWH [10]. While TPT can affect levels of anemia, the impact is not substantial [13].

The disability weights associated with TB among PLWH from GBD 2019 are not distinguished among levels of HIV disease severity. We apply the disability weight associated with TB among PLWH to those in HIV compartment 2 (HIV+, not on ART, CD4 $>$ 200) and TB compartment 6 (active TB). For those in HIV compartment 3 (HIV+, not on ART, CD4 $\leq$ 200) and active TB (in TB compartment 6), we apply the disability weight associated with PLWH without active TB, with a CD4 count of less than 200, with mild anemia to reflect the severity of a CD4 count below 200. For PLWH on ART with active TB (in TB compartment 6 and HIV compartment 4), we assume the same disability as those with active TB who are not living with HIV.

| Parameter Description | Value | Reference |
|---|---|---|
| $D_{t,h}$ Disability weights for those in TB compartment $t$ and HIV compartment $h$, per year | | [16] |
| No active TB and HIV-, $D_{t,1}, t \neq 6$ | 0 (no disability) | |
| No active TB and HIV+, CD4 $>$ 200, $D_{t,2}, t \neq 6$ | 0.016 | |
| No active TB and HIV+, CD4 $\leq$ 200, $D_{t,3}, t \neq 6$ | 0.583 | |
| No active TB and HIV+, on ART, $D_{t,4}, t \neq 6$ | 0.081 | |
| Active TB and HIV-, $D_{6,1}$ | 0.333 | |
| Active TB and HIV+, not on ART, CD4 $>$ 200, $D_{6,2}$ | 0.411 | |
| Active TB and HIV+, not on ART, CD4 $\leq$ 200, $D_{6,3}$ | 0.583 | |
| Active TB and HIV+, on ART, $D_{6,4}$ | 0.333 | |

Table 7: Input parameters values for disability weights used to calculate disability-adjusted life-years (DALYs).

#### 5.6 Parameter Values for Program Costs

The costs in 2018 US dollars are estimated from cited sources for TB and HIV preventative treatments and care as provided in Table 8. These estimates are used to calculate the projected cost of each program using the equations in Section 4.5.

For PLWH on ART, outpatient HIV care costs include the costs associated with administering ART, which is differentiated by delivery method to account for additional costs associated with community-based care versus facility-based care [7]. The inpatient HIV care costs in Table 8

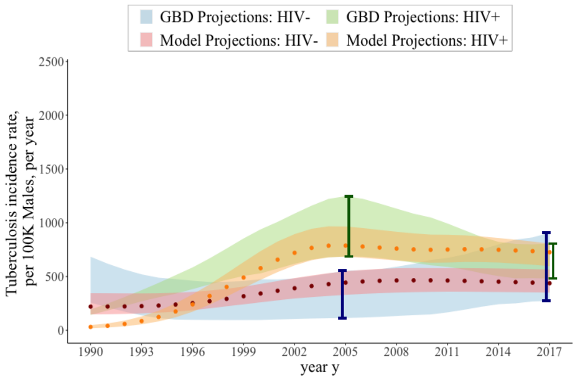

(a) TB incidence rate per 100,000 males in *year y* HIV positive $TBinc\_per_1^{HIV+}(y,1)$ and HIV negative $TBinc\_per_1^{HIV-}(y,1)$

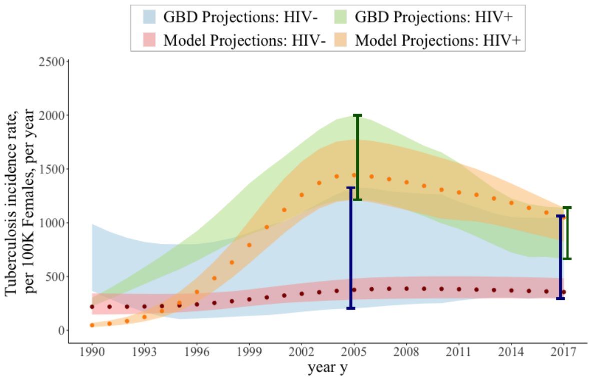

(b) TB incidence rate per 100,000 females in *year y* HIV positive $TBinc\_per_2^{HIV+}(y,1)$ and HIV negative $TBinc\_per_2^{HIV-}(y,1)$

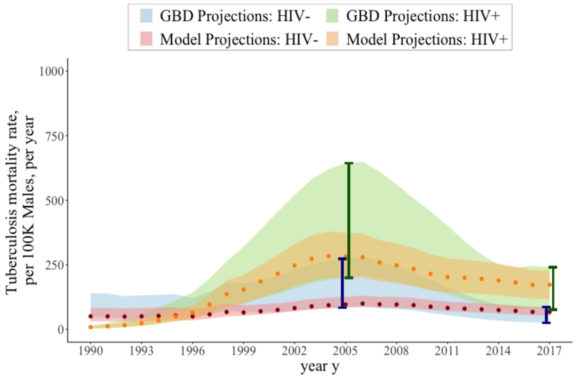

(c) TB mortality rate per 100,000 males in *year y* HIV positive $TBmort\_per_1^{HIV+}(y,1)$ and HIV negative $TBmort\_per_1^{HIV-}(y,1)$

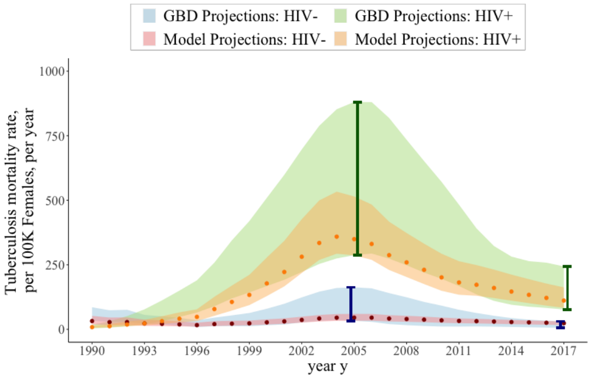

(d) TB mortality rate per 100,000 females in *year y* HIV positive $TBmort\_per_2^{HIV+}(y,1)$ and HIV negative $TBmort\_per_2^{HIV-}(y,1)$

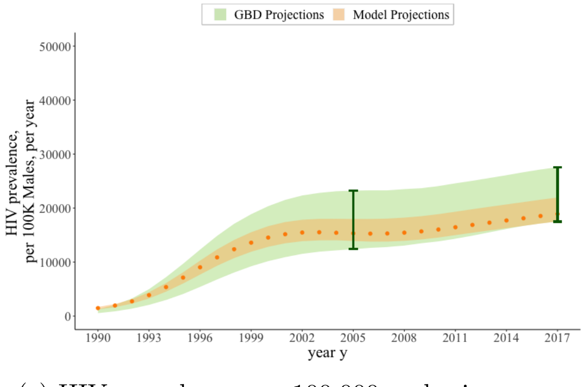

(e) HIV prevalence per 100,000 males in *year y* $HIVprev\_per_1(y,1)$

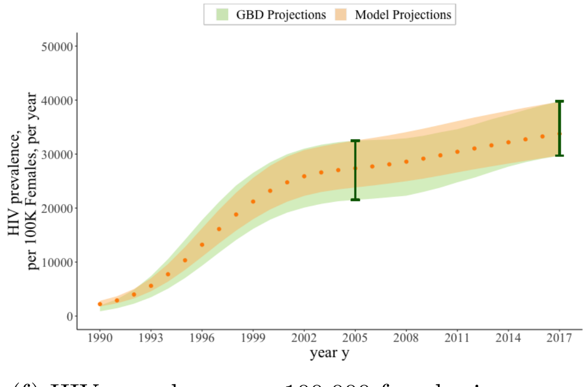

(f) HIV prevalence per 100,000 females in *year y* $HIVprev\_per_2(y,1)$

Figure 5: Maximum, minimum, and mean ranges of TB incidence, TB mortality, and HIV prevalence metrics from the 859 accepted parameter sets for each *year y* under Program 1 ($p = 1$) from 1990 to 2017. Model values are shown in red and orange for HIV negative and HIV positive populations, respectively. Estimates from GBD 2019 for males and females between the ages of 15 and 59 in KwaZulu-Natal, South Africa, are shown in blue and green for HIV negative and HIV positive populations, respectively [4]. In 2005 and 2017, when we compare model values to target calibration ranges, target calibration ranges are emphasized lines.

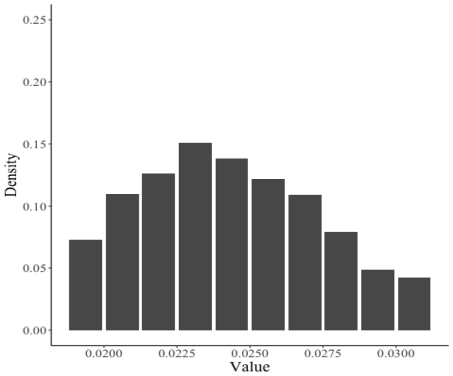

(a) Rate of TB progression from recent LTBI to active TB ($\pi_{3,6}$)

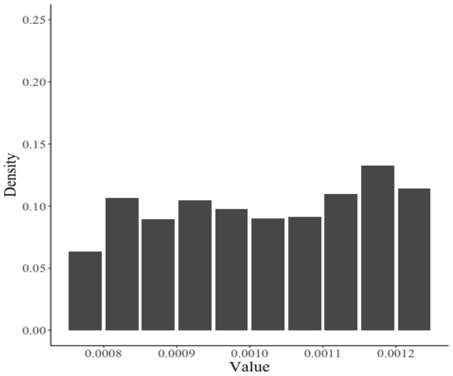

(b) Rate of TB progression from remote LTBI to active TB ($\pi_{4,6}$)

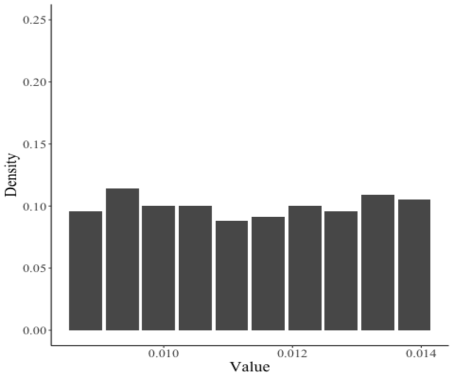

(c) Rate of TB progression from LTBI on TPT to active TB ($\pi_{5,6}$)

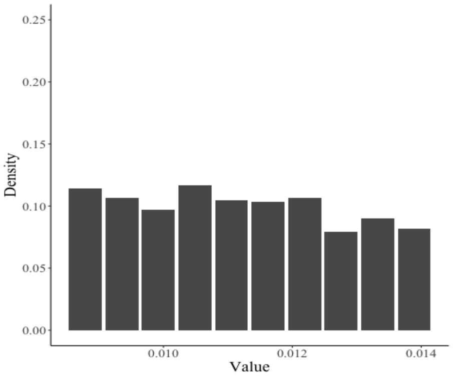

(d) Rate of TB progression from LTBI after TPT to active TB ($\pi_{8,6}$)

Figure 6: Distribution of 859 accepted parameter sets for rates of TB progression from recent LTBI (TB compartment 3), remote LTBI (TB compartment 4), LTBI on TPT (TB compartment 5), and LTBI after TPT (TB compartment 8) to active TB (TB compartment 6).

exclude costs of care for TB-related hospitalization. The total reported HIV inpatient care costs from cited sources include costs of TB-related hospitalization; however, we subtract out the costs of TB-related inpatient care from the total reported HIV inpatient care costs. The estimated HIV testing cost is based on the assumption that programs typically test multiple individuals for HIV before finding an eligible person to initiate ART screening. Based on the screening and enrollment results from the DO ART trial [7], we use a ratio of 5.4 individuals tested for one individual found to be eligible, and the cost per HIV test of \$4.44 [33]. To obtain the cost of HIV testing to find one person to initiate ART, we multiply 5.4 by \$4.44. TPT community-based care is nested in the community-based ART care intervention and only provided to those already on ART, so there are no additional costs associated with community-based TPT care versus facility-based care [7]. TPT costs are defined over a six-month course of TPT and include the cost of the medication (\$8) [43], provider time for counseling (\$12) [8], outpatient care for TPT-associated drug-induced liver injury (DILI) ($6 \times 0.00293 \times \$4$) [23, 38], and provider laboratory costs for TPT-associated DILI ($6 \times 0.00293 \times \$14$) [23, 38], where 0.00293 reflects the probability of developing DILI per month of TPT use. Costs for TB treatment are defined for a course of TB treatment and differentiated by DS-TB and MDR-TB infections to account for the different medication and provider time costs of

| Parameter Description | Value | Reference |
|---|---|---|
| $Out\_Ocare_h^{COST}$ Annual outpatient HIV care costs for PLWH who are: | | |
| Not on ART $Out\_Ocare_h^{COST}, h = \{2,3\}$ | 135 | [18] |
| On ART $Out\_Ocare_h^{COST}, h = 4$ under | | |
| Standard facility-based ART care (Program 1) | 249 | [7] |
| Community-based ART care (Programs 2 and 3) | 310 | [7] |
| $In\_Ocare_h^{COST}$ Annual inpatient HIV care costs for PLWH who are: | | |
| Not on ART, CD4 > 200 $In\_Ocare_2^{COST}$ | 62 | [32] |
| Not on ART, CD4 $\leq$ 200 $In\_Ocare_3^{COST}$ | 162 | [32] |
| On ART (regardless of facility or community-based ART care) $In\_Ocare_4^{COST}$ | 151 | [32] |
| $HIVtest^{COST}$ Cost of HIV testing to find one person to initiate ART | 24 | [7,33] |
| $TPT^{COST}$ Cost of a 6-month course of TPT with isoniazid | 20 | [8,23,38,43] |
| $TBcare_r^{COST}$ Cost of a course of TB treatment for people with | | |
| DS-TB, $TBcare_1^{COST}$ | 259 | [8] |
| MDR-TB, $TBcare_2^{COST}$ | 1,889 | [31] |

Table 8: Input parameter notation and values used in the cost model. Costs are provided in 2018 US dollars. Costs provided in the table are rounded to the nearest dollar.

these treatment regimens [31].

### 6 Supplemental Results

This section provides further detail to supplement the results in the main manuscript's results section.

#### 6.1 TB Incidence and TB Mortality Rates by Gender and HIV status

This section provides TB incidence and TB mortality rates not only by gender (as presented in the main manuscript) but also by HIV status for each of the three care programs over the intervention period from the start of 2018 to the end of 2027. TB incidence rates for PLWH and people without HIV are calculated in Section 4 in Equation (11) and Equation (12), respectively. TB mortality rates for PLWH and people without HIV are calculated in Section 4 using Equation (14) and Equation (15), respectively.

Figure 2 in the manuscript presents TB incidence and TB mortality rates by gender (aggregated over HIV status) for each care program, which is calculated by summing TB incidence and mortality rates for the HIV positive and HIV negative populations for each year and program. In Figure 7 and Figure 8, we present TB incidence and TB mortality rates by gender and HIV status for each of the care programs. Table 9 provides the mean, maximum, and minimum TB incidence and mortality rates in 2027 (the last year of the intervention period) per 100,000 males and females by HIV status and care program over the 859 accepted parameter sets.

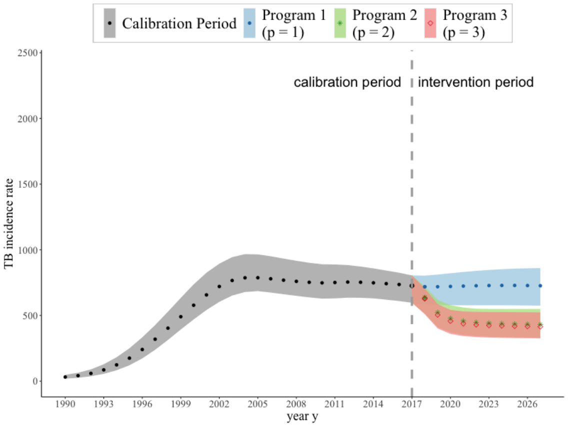

(a) TB incidence per 100,000 males HIV positive $TBinc\_per_1^{HIV+}(y,p)$

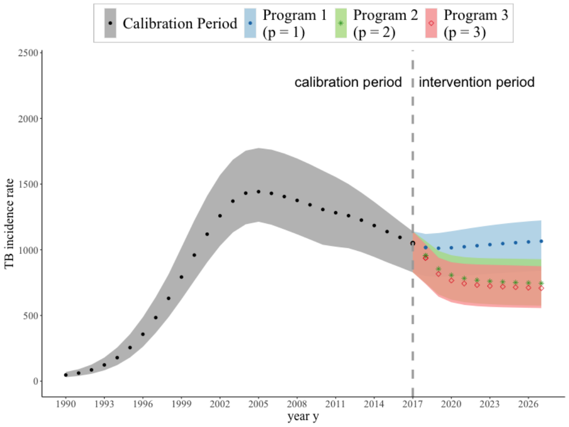

(b) TB incidence per 100,000 females HIV positive $TBinc\_per_2^{HIV+}(y,p)$

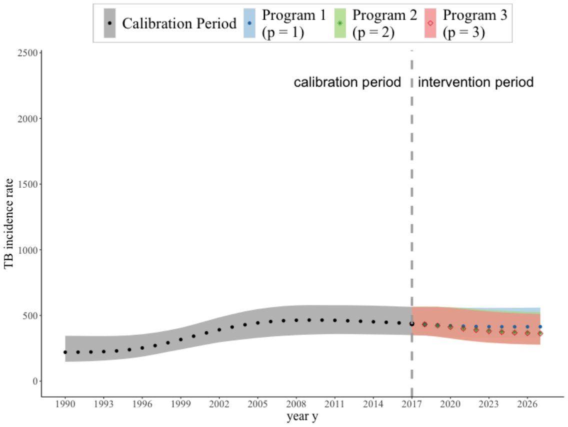

(c) TB incidence per 100,000 males HIV negative $TBinc\_per_1^{HIV-}(y,p)$

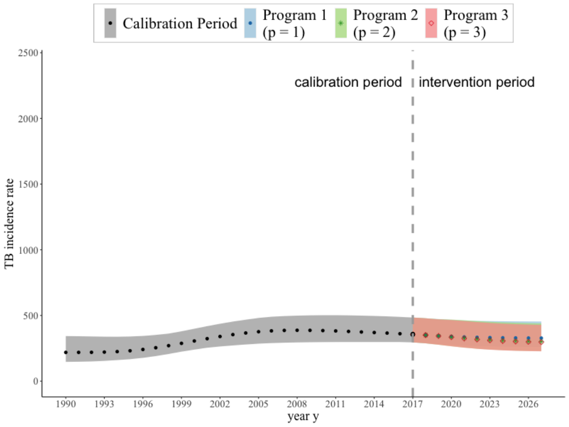

(d) TB incidence per 100,000 females HIV negative $TBinc\_per_2^{HIV-}(y,p)$

Figure 7: Estimated TB incidence by gender in KwaZulu-Natal, South Africa during the calibration period and during the intervention period under the three care programs. Mean, maximum, and minimum yearly TB incidence rates from 1990 to 2017 over the 859 accepted parameter sets are shown in grey (dots). Mean, maximum, and minimum yearly TB incidence rates during the intervention period (2018-2027) over the 859 accepted parameter sets are illustrated by care program. During the intervention period, Program 1 (standard facility-based ART and TPT care) is shown in blue (dots), Program 2 (community-based ART care with standard facility-based TPT) is shown in green (stars), and Program 3 (community-based ART and TPT care) is shown in red (diamonds).

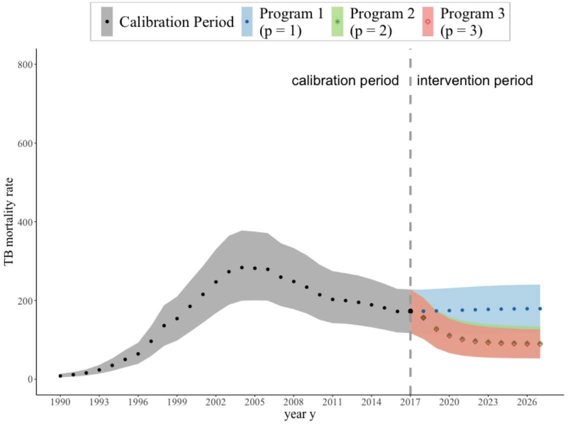

(a) TB mortality per 100,000 males and HIV positive $TBmort\_per_1^{HIV+}(y,p)$

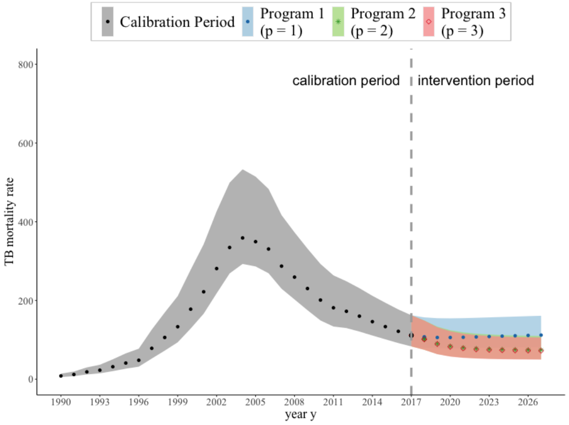

(b) TB mortality per 100,000 females and HIV positive $TBmort\_per_2^{HIV+}(y,p)$

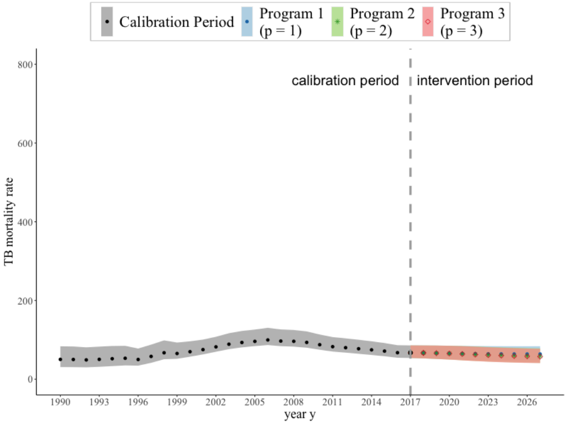

(c) TB mortality per 100,000 males and HIV negative $TBmort\_per_1^{HIV-}(y,p)$

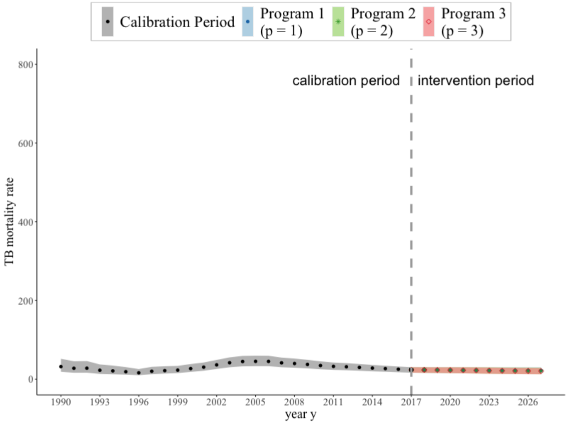

(d) TB mortality per 100,000 females and HIV negative $TBmort\_per_2^{HIV-}(y,p)$

Figure 8: Estimated TB mortality rates by gender in KwaZulu-Natal, South Africa during the calibration period and during the intervention period under the three ART and TPT care programs. Mean, maximum, and minimum yearly TB mortality rates from 1990 to 2017 over the 859 accepted parameter sets are shown in grey (dots). Mean, maximum, and minimum yearly TB mortality rates during the intervention period (2018-2027) over the 859 accepted parameter sets are illustrated by care program. During the intervention period, Program 1 (standard facility-based ART and TPT care) is shown in blue (dots), Program 2 (community-based ART care with standard facility-based TPT) is shown in green (stars), and Program 3 (community-based ART and TPT care) is shown in red (diamonds).

| | **Program 1** *Standard facility-based ART and TPT care* $(p = 1)$ | **Program 2** *Community-based ART care with standard facility-based TPT care* $(p = 2)$ | **Program 3** *Community-based ART with TPT care* $(p = 3)$ |
|---|---|---|---|
| | **Mean [Min, Max]** | **Mean [Min, Max]** | **Mean [Min, Max]** |
| **TB incidence rate** | | | |
| TB incidence rate per 100,000 males and HIV positive $TBinc\_per_1^{HIV+}(2027,\ p)$ | 727 [609, 818] | 432 [358, 515] | 414 [345, 492] |
| TB incidence rate per 100,000 females and HIV positive $TBinc\_per_2^{HIV+}(2027,\ p)$ | 1066 [899, 1184] | 745 [613, 863] | 707 [586, 819] |
| TB incidence rate per 100,000 males and HIV negative $TBinc\_per_1^{HIV-}(2027,\ p)$ | 414 [340, 504] | 367 [296, 456] | 361 [293, 451] |
| TB incidence rate per 100,000 females and HIV negative $TBinc\_per_2^{HIV-}(2027,\ p)$ | 328 [269, 413] | 301 [238, 388] | 296 [235, 374] |
| **TB mortality rate** | | | |
| TB mortality rate per 100,000 males and HIV positive $TBmort\_per_1^{HIV+}(2027,\ p)$ | 179 [131, 230] | 91 [62, 122] | 89 [62, 117] |
| TB mortality rate per 100,000 females and HIV positive $TBmort\_per_2^{HIV+}(2027,\ p)$ | 112 [86, 150] | 74 [55, 99] | 72 [53, 97] |
| TB mortality rate per 100,000 males and HIV negative $TBmort\_per_1^{HIV-}(2027,\ p)$ | 64 [51, 81] | 58 [44, 75] | 57 [44, 74] |
| TB mortality rate per 100,000 females and HIV negative $TBmort\_per_2^{HIV-}(2027,\ p)$ | 22 [15, 29] | 21 [14, 28] | 21 [14, 27] |

Table 9: Mean, minimum, and maximum TB incidence and mortality rates in 2027 (the last year of the intervention period) per 100,000 males and 100,000 females by HIV status and care program over the 859 accepted parameter sets.

#### 6.2 IPT Initiation under Facility-Based and Community-Based TPT Care Programs Over Time

Figure 9 illustrates the average number of individuals initiating TPT from the start of 2018 to the end of 2027 for facility-based TPT care programs, including Program 1 (standard facility-based ART and TPT care) and Program 2 (community-based ART care with standard facility-based TPT care), and community-based TPT care, Program 3 (community-based ART with TPT). Under Program 3 (community-based ART with TPT care), we model a rapid TPT scale-up to reach the initiation rates observed in the DO ART trial, which would result in many individuals taking TPT within the first few years of the intervention. Over time, maintaining the same TPT initiation rates among eligible PLWH on ART would result in a decline in the number of people on TPT, as PLWH on ART would have already completed a course of TPT. Over the 10-year intervention period, an estimated 31,009 (range 25,674 – 39,021) PLWH received a course of TPT in Program 3 compared to 17,264 (range 14,156 – 21,255) in Program 1 and 18,221 (range 15,305 – 22,370) in Program 2.

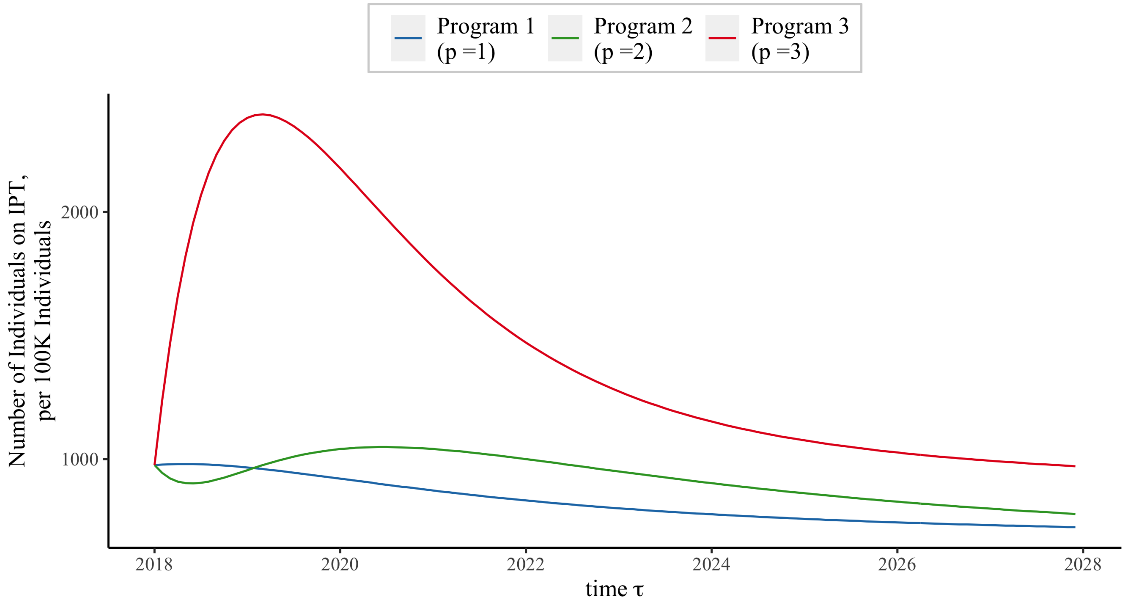

Figure 9: Mean number of individuals on TPT per 100,000 individuals over all 859 accepted calibration sets overtime during the intervention period from the start of 2018 to the end of 2027 by care program. Program 1 (standard facility-based ART and TPT care), Program 2 (community-based ART care with standard facility-based TPT), and Program 3 (community-based ART and TPT care) are highlighted in blue, green, and red, respectively.

[11] Connolly C, Reid A, Davies G, Sturm W, McAdam KPWJ, Wilkinson D. Relapse and mortality among HIV-infected and uninfected patients with tuberculosis successfully treated with twice weekly directly observed therapy in rural South Africa. AIDS [Internet].

1999;13(12). Available from:
https://journals.lww.com/aidsonline/Fulltext/1999/08200/Relapse_and_mortality_among_HIV_infected_and.1

[12] Dale KD, Karmakar M, Snow KJ, Menzies D, Trauer JM, Denholm JT. Quantifying the rates of late reactivation tuberculosis: a systematic review. Lancet Infect Dis [Internet]. 2021; Available from: https://www.sciencedirect.com/science/article/pii/S1473309920307283

[13] Danel C, Moh R, Gabillard D, Badje A, Le Carrou J, Ouassa T, et al. A trial of early antiretrovirals and isoniazid preventive therapy in Africa. N Engl J Med. 2015;373(9).

[14] Egsmose T, Ang'awa JO, Poti SJ. The use of isoniazid among household contacts of open cases of pulmonary tuberculosis. Bull World Health Organization [Internet]. 1965;33(3):419–33. Available from: http://www.ncbi.nlm.nih.gov/pubmed/5321762

[15] Ferebee SH, Mount FW. Tuberculosis Morbidity in a Controlled Trial of the Prophylactic Use of Isoniazid among Household Contacts. Am Rev Respir Dis [Internet]. 1962 Apr 1;85(4):490–510. Available from: https://www.atsjournals.org/doi/abs/10.1164/arrd.1962.85.4.490

[16] Global Burden of Disease Collaborative Network. Global Burden of Disease Study 2019 (GBD2019) Disability Weights. 2020.

[50] World Health Organization. Global Tuberculosis Report. Geneva, Switzerland; 2020.
